# Design and methodological considerations for biomarker discovery and validation in the Integrative Analysis of Lung Cancer Etiology and Risk (INTEGRAL) Program

**DOI:** 10.1101/2022.03.21.22272544

**Authors:** Hilary A Robbins, Karine Alcala, Elham Khodayari Moez, Florence Guida, Sera Thomas, Hana Zahed, Matthew T Warkentin, Karl Smith-Byrne, Yonathan Brhane, David Muller, Demetrius Albanes, Melinda C Aldrich, Alan A Arslan, Julie Bassett, Christine D Berg, Qiuyin Cai, Chu Chen, Michael PA Davies, Brenda Diergaarde, John K Field, Neal D Freedman, Wen-Yi Huang, Mikael Johansson, Michael Jones, Woon-Puay Koh, Stephen Lam, Qing Lan, Arnulf Langhammer, Linda M Liao, Geoffrey Liu, Reza Malekzadeh, Roger L Milne, Luis M Montuenga, Thomas Rohan, Howard D Sesso, Gianluca Severi, Mahdi Sheikh, Rashmi Sinha, Xiao-Ou Shu, Victoria L Stevens, Martin C Tammemägi, Lesley F Tinker, Kala Visvanathan, Ying Wang, Renwei Wang, Stephanie J Weinstein, Emily White, David Wilson, Jian-Min Yuan, Xuehong Zhang, Wei Zheng, Christopher I Amos, Paul Brennan, Mattias Johansson, Rayjean J Hung

**Author notes:** Contributed equally (KA, EKM). Joint senior authors (MJ, RJH). **Corresponding authors:** Hilary Robbins and Mattias Johansson, Genomic Epidemiology Branch, International Agency for Research on Cancer, 150 cours Albert Thomas, CEDEX 69732 Lyon, France, Rayjean Hung, Lunenfeld-Tanenbaum Research Institute, Sinai Health, Dalla Lana School of Public Health, University of Toronto, 60 Murray St. Toronto, ON M5T 3L9. Canada. **Disclaimer:** Where authors are identified as personnel of the International Agency for Research on Cancer / World Health Organization, the authors alone are responsible for the views expressed in this article and they do not necessarily represent the decisions, policy, or views of the International Agency for Research on Cancer / World Health Organization. The contents of this manuscript are solely the responsibility of the authors and do not necessarily represent the official views of the Centers for Disease Control and Prevention or the Department of Health and Human Services, nor does mention of trade names, commercial products, or organizations imply endorsement by the US government.

## Abstract

The Integrative Analysis of Lung Cancer Etiology and Risk (INTEGRAL) program is an NCI-funded initiative with an objective to develop tools to optimize lung cancer screening. Here, we describe the rationale and design for the Risk Biomarker and Nodule Malignancy projects within INTEGRAL.

The overarching goal of these projects is to systematically investigate circulating protein markers to include on a panel for use (i) pre-LDCT, to identify people likely to benefit from screening, and (ii) post-LDCT, to differentiate benign versus malignant nodules. To identify informative proteins, the Risk Biomarker project measured 1,161 proteins in a nested-case control study within 2 prospective cohorts (n=252 lung cancer cases and 252 controls) and replicated associations for a subset of proteins in 4 cohorts (n=479 cases and 479 controls). Eligible participants had any history of smoking and cases were diagnosed within 3 years of blood draw. The Nodule Malignancy project measured 1,077 proteins among participants with a heavy smoking history within 4 LDCT screening studies (n=425 cases within 5 years of blood draw, 398 benign-nodule controls, and 430 nodule-free controls).

The INTEGRAL panel will enable absolute quantification of 21 proteins. We will evaluate its lung cancer discriminative performance in the Risk Biomarker project using a case-cohort study including 14 cohorts (n=1,696 cases and 2,926 subcohort representatives), and in the Nodule Malignancy project within 5 LDCT screening studies (n=675 cases, 648 benign-nodule controls, and 680 nodule-free controls). Future progress to advance lung cancer early detection biomarkers will require carefully designed validation, translational, and comparative studies.

## Introduction

Lung cancer screening by low-dose computed tomography (LDCT) has accelerated the field of lung cancer research with a renewed focus on early detection.^1,2^ However, several questions remain regarding how to best implement LDCT screening,^3^ including how to identify individuals who are likely to benefit from screening, and how to manage nodules of indeterminate malignancy status identified on LDCT scans.

In 2018, the US National Cancer Institute (NCI) funded the Integrative Analysis of Cancer Risk and Etiology (INTEGRAL) U19 program, which includes an objective to develop early detection biomarkers and risk prediction tools for lung cancer screening. The INTEGRAL program comprises 3 projects: the Genetics project focused on germline genetics, the Risk Biomarker project focused on pre-diagnostic blood biomarkers, and the Nodule Malignancy project focused on applications in LDCT screening studies including nodule evaluation. Here, we describe a joint effort of the Risk Biomarker and Nodule Malignancy projects to systematically investigate circulating protein markers for both pre- and post-LDCT applications.

The primary objective of the Risk Biomarker project is to identify and validate biomarkers that can improve lung cancer risk prediction among people with a smoking history. A secondary objective is to develop and validate questionnaire-based lung cancer risk prediction models. The objectives for the Nodule Malignancy project are to identify biomarkers and establish quantitative imaging models that can differentiate benign versus malignant nodules following an initial LDCT scan. The Risk Biomarker project leverages resources from the Lung Cancer Cohort Consortium (LC3)^4–8^ which was initially established in 2010 within the NCI Cohort Consortium.^9^ The Nodule Malignancy project brings together LDCT screening studies in the framework of the International Lung Cancer Consortium (ILCCO), which has provided a foundation for collaborative research on lung cancer since 2004 (http://ilcco.iarc.fr).

Herein, we provide a design overview of the biomarker studies within the INTEGRAL Risk Biomarker and Nodule Malignancy projects. We highlight considerations that motivated the design, present details of the study population, and describe the harmonized databases resulting from these projects. Finally, we discuss perspectives for research to follow this initiative with a view toward implementation of the prediction tools in clinical practice.

### Development and validation of a protein biomarker panel for early lung cancer detection

#### Motivation

The US Preventive Services Task Force (USPSTF) currently recommends lung cancer screening for people aged 50-80 years who have smoked at least 20 pack-years and currently smoke or have quit within the past 15 years.^10^ However, more than one-third of lung cancer deaths that could be prevented among people who have smoked fall outside of these criteria.^11^ To better target the highest-risk population, screening can instead be offered to people whose individual lung cancer risk exceeds a certain threshold as estimated by a risk prediction model.^12–15^ This approach is included in the US National Comprehensive Cancer Network (NCCN) guidelines.^16^

Biomarkers may provide additional or complementary information on lung cancer risk and represent a promising avenue to improve existing risk prediction models. Conceptually, this could improve efficiency in two ways: by offering screening to people who have high risk based on biomarkers but are not otherwise eligible for screening based on the current recommendation, and by deprioritizing screening for individuals who are eligible but have a low-risk biomarker profile. Various domains of biomarkers have been investigated, but the translation of this research into practice has been slow, partly due to the lack of appropriately designed studies to establish and validate biomarker-based risk prediction models.^17,18^

Another setting in which biomarkers could be applied in lung cancer screening is to better distinguish between malignant and benign nodules on LDCT images. Nodules are detected in up to one-quarter of participants, but the vast majority are benign. Managing nodules with uncertain clinical significance (i.e., indeterminate nodules) represents an important challenge because false-positive nodules can lead to interventions with risks of long-term harm. On the other hand, missed malignant nodules can lead to a lost opportunity for curative treatment. Several prediction models for nodule malignancy have been developed,^19–21^ but their classification accuracies remain imperfect.

Recent papers have highlighted common limitations in the design of studies aiming to identify and validate biomarkers for early cancer detection^22^ including lung cancer.^18^ To avoid common biases resulting from systematic differences between cases and controls, the prospective-specimen-collection, retrospective-blinded-evaluation (PRoBE) design emphasizes the use of pre-diagnostic samples, sampling from the same source population, and matching on important factors that impact biomarker measurements and outcome.^23^ In validation studies, it is critical that the added contribution of the biomarker, compared with existing tools, can be clearly identified and quantified.^18^

In a pilot study published in 2018, members of our team found that a pre-defined set of cancer-related protein biomarkers improved discrimination between lung cancer cases and controls compared to a smoking-based risk prediction model, when the markers were measured in an independent validation study using samples collected within the year before diagnosis.^24^ Studies also suggest that protein markers can improve discrimination between malignant and benign lung nodules.^25,26^ Building on these promising preliminary data, the INTEGRAL program was formed to conduct a comprehensive protein biomarker evaluation from discovery to validation for both population-based risk prediction (Risk Biomarker project), and nodule differentiation (Nodule Malignancy project).

Our overarching aims are *i)* to identify circulating proteins that provide additional information to the gold standard on both lung cancer risk and nodule malignancy and *ii)* to develop and validate a multiplex lung cancer biomarker assay that can quantify key lung cancer risk and/or nodule-malignancy proteins in small volumes of peripheral blood in a cost-effective manner. Use of a single assay will help to streamline clinical implementation along the various steps of the LDCT screening pathway.

#### Design

##### Overview

**Figure 1** outlines the sequential study phases of the INTEGRAL Risk Biomarker and Nodule Malignancy projects. In the Risk Biomarker project, an initial ‘full discovery’ phase scanned a broad set of protein markers, followed by a ‘targeted discovery’ phase which replicated results for a subset of proteins. The Nodule Malignancy project started with an expanded targeted discovery phase and analyzed samples from LDCT screening studies to identify proteins that are specifically useful to distinguish between benign and malignant lung nodules. The results from both projects will be used to configure the INTEGRAL panel with 21 circulating protein markers, whose performance will be assessed in a validation phase. **Table 1** summarizes the key characteristics of the participating cohorts and LDCT screening studies in each phase.

**Figure 1:**
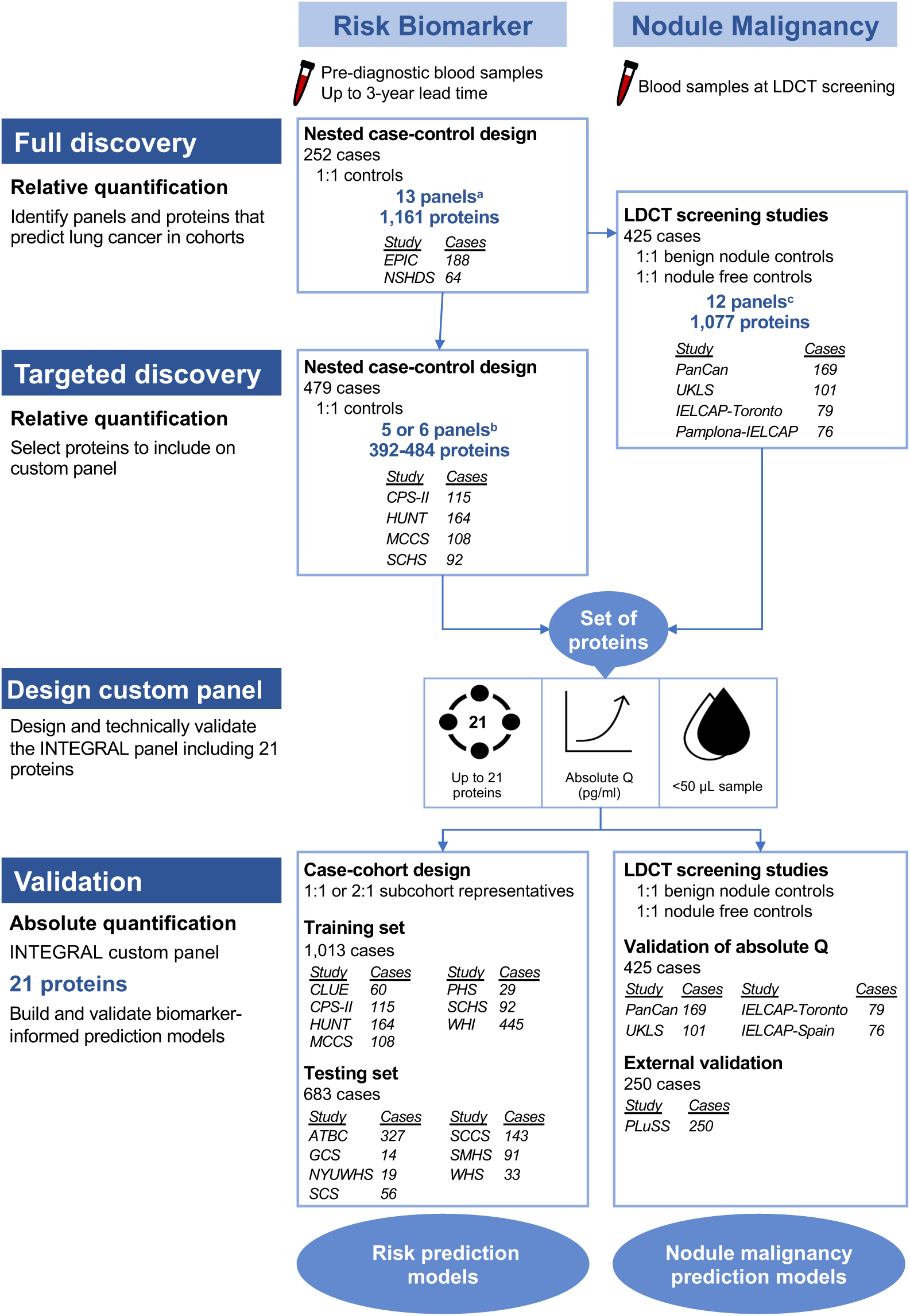
Schematic describing the development and validation of the INTEGRAL protein panel for lung cancer early detection and nodule malignancy. See Table 1 for definitions of the cohort abbreviations. a: Cardiometabolic, Cardiovascular II, Cardiovascular III, Cell Regulation, Development, Immune response, Inflammation, Metabolism, Neurology Oncology II, Oncology III, Organ Damage, NeuroExploratory b: Cardiovascular III, Inflammation, Immuno-Oncology, Oncology II, Oncology III, NeuroExploratory c: Cardiometabolic, Cardiovascular II, Cardiovascular III, Development, Immune Response, Inflammation, Metabolism, Neurology Oncology II, Oncology III, Organ Damage, NeuroExploratory

**Table 1:**
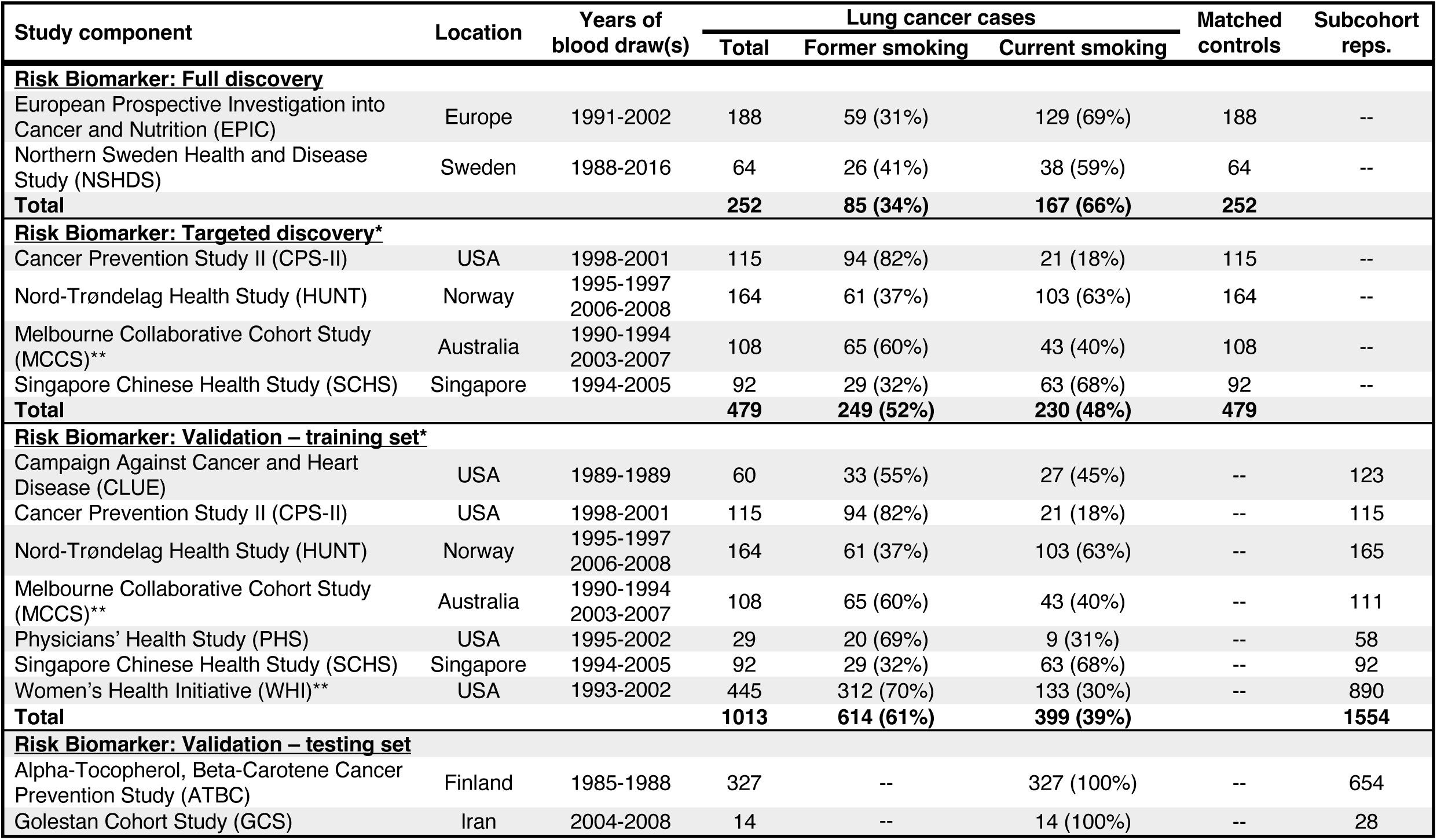
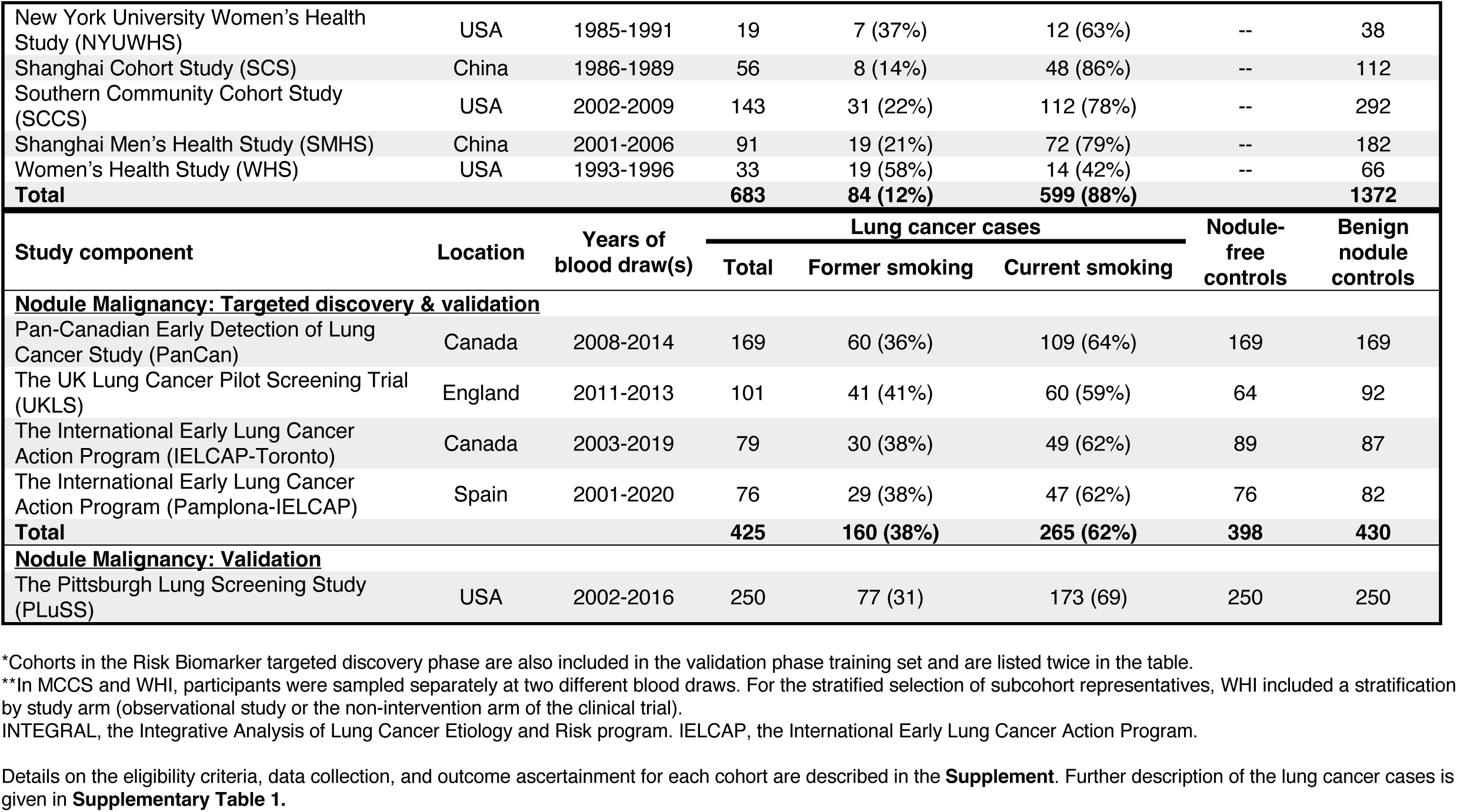
Description of lung cancer cases participating in the development and validation of the INTEGRAL protein panel for lung cancer early detection and nodule malignancy.

| Study component | Location | Years of blood draw(s) | Lung cancer cases |  |  | Matched controls | Subcohort reps. |
| --- | --- | --- | --- | --- | --- | --- | --- |
|  |  |  | Total | Former smoking | Current smoking |  |  |
| **<u>Risk Biomarker: Full discovery</u>** |  |  |  |  |  |  |  |
| European Prospective Investigation into Cancer and Nutrition (EPIC) | Europe | 1991-2002 | 188 | 59 (31%) | 129 (69%) | 188 | -- |
| Northern Sweden Health and Disease Study (NSHDS) | Sweden | 1988-2016 | 64 | 26 (41%) | 38 (59%) | 64 | -- |
| **Total** |  |  | **252** | **85 (34%)** | **167 (66%)** | **252** |  |
| **<u>Risk Biomarker: Targeted discovery\*</u>** |  |  |  |  |  |  |  |
| Cancer Prevention Study II (CPS-II) | USA | 1998-2001 | 115 | 94 (82%) | 21 (18%) | 115 | -- |
| Nord-Trøndelag Health Study (HUNT) | Norway | 1995-1997<br>2006-2008 | 164 | 61 (37%) | 103 (63%) | 164 | -- |
| Melbourne Collaborative Cohort Study (MCCS)** | Australia | 1990-1994<br>2003-2007 | 108 | 65 (60%) | 43 (40%) | 108 | -- |
| Singapore Chinese Health Study (SCHS) | Singapore | 1994-2005 | 92 | 29 (32%) | 63 (68%) | 92 | -- |
| **Total** |  |  | **479** | **249 (52%)** | **230 (48%)** | **479** |  |
| **<u>Risk Biomarker: Validation – training set\*</u>** |  |  |  |  |  |  |  |
| Campaign Against Cancer and Heart Disease (CLUE) | USA | 1989-1989 | 60 | 33 (55%) | 27 (45%) | -- | 123 |
| Cancer Prevention Study II (CPS-II) | USA | 1998-2001 | 115 | 94 (82%) | 21 (18%) | -- | 115 |
| Nord-Trøndelag Health Study (HUNT) | Norway | 1995-1997<br>2006-2008 | 164 | 61 (37%) | 103 (63%) | -- | 165 |
| Melbourne Collaborative Cohort Study (MCCS)** | Australia | 1990-1994<br>2003-2007 | 108 | 65 (60%) | 43 (40%) | -- | 111 |
| Physicians' Health Study (PHS) | USA | 1995-2002 | 29 | 20 (69%) | 9 (31%) | -- | 58 |
| Singapore Chinese Health Study (SCHS) | Singapore | 1994-2005 | 92 | 29 (32%) | 63 (68%) | -- | 92 |
| Women's Health Initiative (WHI)** | USA | 1993-2002 | 445 | 312 (70%) | 133 (30%) | -- | 890 |
| **Total** |  |  | **1013** | **614 (61%)** | **399 (39%)** |  | **1554** |
| **<u>Risk Biomarker: Validation – testing set</u>** |  |  |  |  |  |  |  |
| Alpha-Tocopherol, Beta-Carotene Cancer Prevention Study (ATBC) | Finland | 1985-1988 | 327 | -- | 327 (100%) | -- | 654 |
| Golestan Cohort Study (GCS) | Iran | 2004-2008 | 14 | -- | 14 (100%) | -- | 28 |

| New York University Women’s Health Study (NYUWHS) | USA | 1985-1991 | 19 | 7 (37%) | 12 (63%) | -- | 38 |
| --- | --- | --- | --- | --- | --- | --- | --- |
| Shanghai Cohort Study (SCS) | China | 1986-1989 | 56 | 8 (14%) | 48 (86%) | -- | 112 |
| Southern Community Cohort Study (SCCS) | USA | 2002-2009 | 143 | 31 (22%) | 112 (78%) | -- | 292 |
| Shanghai Men’s Health Study (SMHS) | China | 2001-2006 | 91 | 19 (21%) | 72 (79%) | -- | 182 |
| Women’s Health Study (WHS) | USA | 1993-1996 | 33 | 19 (58%) | 14 (42%) | -- | 66 |
| Total |  |  | 683 | 84 (12%) | 599 (88%) |  | 1372 |
| Study component | Location | Years of blood draw(s) | Lung cancer cases |  |  | Nodule-free controls | Benign nodule controls |
|  |  |  | Total | Former smoking | Current smoking |  |  |
| **<u>Nodule Malignancy: Targeted discovery & validation</u>** |  |  |  |  |  |  |  |
| Pan-Canadian Early Detection of Lung Cancer Study (PanCan) | Canada | 2008-2014 | 169 | 60 (36%) | 109 (64%) | 169 | 169 |
| The UK Lung Cancer Pilot Screening Trial (UKLS) | England | 2011-2013 | 101 | 41 (41%) | 60 (59%) | 64 | 92 |
| The International Early Lung Cancer Action Program (IELCAP-Toronto) | Canada | 2003-2019 | 79 | 30 (38%) | 49 (62%) | 89 | 87 |
| The International Early Lung Cancer Action Program (Pamplona-IELCAP) | Spain | 2001-2020 | 76 | 29 (38%) | 47 (62%) | 76 | 82 |
| Total |  |  | 425 | 160 (38%) | 265 (62%) | 398 | 430 |
| **<u>Nodule Malignancy: Validation</u>** |  |  |  |  |  |  |  |
| The Pittsburgh Lung Screening Study (PLuSS) | USA | 2002-2016 | 250 | 77 (31) | 173 (69) | 250 | 250 |

\*Cohorts in the Risk Biomarker targeted discovery phase are also included in the validation phase training set and are listed twice in the table.
\*\*In MCCS and WHI, participants were sampled separately at two different blood draws. For the stratified selection of subcohort representatives, WHI included a stratification by study arm (observational study or the non-intervention arm of the clinical trial).
INTEGRAL, the Integrative Analysis of Lung Cancer Etiology and Risk program. IELCAP, the International Early Lung Cancer Action Program.

We are using the Olink proteomics platform (Olink Proteomics, Uppsala, Sweden) throughout the project.^27^ Olink discovery assays allow high-throughput semi-quantified concentration measures of highly annotated proteins in less than 50 uL of plasma or serum. The technology uses a proximity extension assay (PEA) technique that is highly sensitive and avoids cross-reactivity, with high reproducibility. Relative protein concentrations are expressed as normalized protein expression (NPX) on log2 scale, which is estimated from quantitative PCR cycle threshold values.

To enable absolute quantification of proteins for clinical applications, we will develop the INTEGRAL panel at Olink. Olink customized panels are also based on PEA technology and can measure up to 21 proteins in less than 50 uL of plasma or serum.^28^ We plan to include 21 proteins on our panel since reducing the number of proteins reduces neither the assay cost nor the sample volume requirement. For all laboratory analyses, cases and controls were randomly allocated over the 96-well plates, with matched pairs plated together where relevant.

##### Risk Biomarker project

The design of the Risk Biomarker project was informed by several considerations. First, we restricted to participants who currently or formerly smoked because they represent the current target population for lung cancer screening.^10^ Second, we included cases diagnosed within 3 years following blood draw, to predict lung cancer within a clinically actionable timeframe.^24^ Third, we used a matched case-control design for the discovery phases, but a case-cohort design for the validation phase. For discovery, the matched design is important to eliminate influences such as storage duration and biospecimen handling. In the validation phase, we changed to a case-cohort design, where the controls were randomly selected from each cohort, to facilitate development of an integrated risk prediction model that is well-calibrated and representative of the source population (i.e., the full cohorts).

###### Full discovery phase

In the Risk Biomarker project full discovery phase, we measured all 13 Olink proteomics panels available in late 2019, which cover a range of domains including inflammation, oncology, and cardiovascular disease (1,161 proteins, **Appendix Spreadsheet, Table 2**). The objective of the full discovery phase was to select panels to measure in the targeted discovery phase, and the sample included the European Investigation into Cancer and Nutrition (EPIC, n=188 lung cancer cases) and the Northern Sweden Health and Disease Study (NSHDS, n=64 cases) (**Table 1;** further details in **Supplementary Table 1**). We included all confirmed lung cancer cases among people who ever smoked that were diagnosed within 3 years of blood draw. For each case, one control was randomly chosen using incidence density sampling from risk sets consisting of people who ever smoked and were alive and free of cancer at the time of diagnosis of the index case. Matching criteria included cohort, study center (where relevant), sex, date of blood collection (±1 month, relaxed to ±3 months for sets without available controls), date of birth (±1 year, relaxed to ±3 years), and smoking status in 4 categories: people who formerly smoked and quit <10 or ≥10 years prior, and people who currently smoked <15 or ≥15 cigarettes per day.

**Table 2:** Proteomics panels tested in the full and targeted discovery phases to develop the INTEGRAL protein panel for lung cancer early detection and nodule malignancy.

| Cohorts | Risk Biomarker Project |  |  |  |  |  | Nodule Malignancy Project |  |  |  |
| --- | --- | --- | --- | --- | --- | --- | --- | --- | --- | --- |
|  | Full Discovery |  | Targeted Discovery |  |  |  | Targeted Discovery |  |  |  |
|  | EPIC | NSHDS | SCHS | CPS-II | HUNT | MCCS | PanCan | UKLS | IELCAP-Toronto | Pamplona-IELCAP |
| Number of cases | 188 | 64 | 92 | 115 | 163 | 108 | 169 | 101 | 79 | 76 |
| Number of panels measured | 13 | 13 | 5 | 6 | 5 | 6 | 12 | 12 | 12 | 12 |
| Number of Olink IDs* | 1196 | 1196 | 460 | 552 | 460 | 552 | 1104 | 1104 | 1104 | 1104 |
| Number of unique proteins* | 1161 | 1161 | 394 | 484 | 392 | 484 | 1077 | 1077 | 1077 | 1077 |
| Proteomics panels |  |  |  |  |  |  |  |  |  |  |
| Cardiovascular III | X | X | X | X | X | X | X | X | X | X |
| Inflammation | X | X | X | X | X | X | X | X | X | X |
| Immuno-Oncology | (X) | (X) | X | X | X | X | (X) | (X) | (X) | (X) |
| Oncology II | X | X | X | X | X | X | X | X | X | X |
| Oncology III | X | X | X | X |  | X | X | X | X | X |
| NeuroExploratory | X | X |  | X | X | X | X | X | X | X |
| Cardiometabolic | X | X |  |  |  |  | X | X | X | X |
| Cardiovascular II | X | X |  |  |  |  | X | X | X | X |
| Cell Regulation | X | X |  |  |  |  |  |  |  |  |
| Development | X | X |  |  |  |  | X | X | X | X |
| Immune Response | X | X |  |  |  |  | X | X | X | X |
| Metabolism | X | X |  |  |  |  | X | X | X | X |
| Neurology | X | X |  |  |  |  | X | X | X | X |
| Organ Damage | X | X |  |  |  |  | X | X | X | X |

\*Some proteins are measured on multiple panels and therefore have multiple Olink IDs for the same protein. In these cases, for each protein, we chose a single Olink ID for analysis by choosing the one that was measured on more cohorts, and then if needed, the Olink ID with the highest variance.
(X): all the proteins from the Immuno-Oncology panel are included on other panels assayed as indicated.
Details of the proteins measured on each panel are provided in the Appendix Table.

The dataset generated by the full discovery phase therefore included 252 case-control pairs with 1,161 proteins measured on each participant (**Table 2**). Statistical analyses applied conditional logistic and penalized regression. We used the results to examine, for each of the 13 proteomics panels, the number of highly ranked and consistently selected proteins.

###### Targeted discovery phase

The targeted discovery phase of the Risk Biomarker project used the same design to independently replicate associations for a subset of proteomics panels, chosen to maximize coverage of the promising proteins while minimizing the total cost. This phase included 4 cohorts: the Cancer Prevention Study II, the Nord-Trøndelag Health Study, the Melbourne Collaborative Cohort Study, and the Singapore Chinese Health Study (**Table 1;** further details in **Supplementary Table 1**). We measured the Immuno-oncology, Oncology II, Cardiovascular III, and Inflammation panels on all 4 cohorts, and the Oncology III and Neuro-exploratory panels on 3 cohorts each (**Table 2**).

The dataset generated for the targeted discovery phase therefore included 479 case-control pairs with between 392 and 484 proteins measured for each participant (**Table 2**). Statistical analyses included conditional logistic regression, penalized regression, and stratified approaches. For the INTEGRAL panel, we are prioritizing proteins selected in penalized regression models that show a consistent association with lung cancer across cohorts.

###### Validation phase

The Risk Biomarker project validation phase employs a case-cohort design including cases diagnosed within 3 years of blood draw. Subcohort representatives were randomly sampled at the time of blood draw in 8 jointly defined categories including age (above or below the median age among cases), sex (male or female, except for single-sex cohorts), and smoking status (current or former). The full baseline cohorts of participants who ever smoked can then be easily represented by inverse-probability weighting. To maximize statistical power, we included the 4 cohorts from the targeted discovery phase again in the validation phase, with 1 subcohort representative per case. The 10 cohorts included only in the validation phase contributed 2 representatives per case.

The optimization process for the INTEGRAL panel is currently underway. Once complete, the validation phase samples will be assayed for absolute quantification of the 21 proteins on the INTEGRAL panel. The cohorts will be divided into training and testing sets (**Table 1**). To maintain full independence of the testing set, the 4 cohorts that contributed to the targeted discovery phase will be included in the training set only. The training set will additionally include the Campaign Against Cancer and Heart Disease, the Physicians’ Health Study, and the Women’s Health Initiative. The testing set will include the Alpha-Tocopherol, Beta-Carotene Cancer Prevention Study, the Golestan Cohort Study, the New York University Women’s Health Study, the Shanghai Cohort Study, the Southern Community Cohort Study, the Shanghai Men’s Health Study, and the Women’s Health Study. These groupings were chosen to balance the training and testing sets by geographical location, US racial/ethnic groups, people who currently or formerly smoked, and lung cancer histological types.

Statistical analyses in the validation phase will use the training set to establish flexible parametric survival models that predict absolute risk of lung cancer over 3 years.^29^ Predictors will include a subset of the 21 proteins from the INTEGRAL panel in addition to demographic, health history, and smoking information. The final model will be evaluated in the testing set to measure its calibration (ratio of observed to expected cases) and discrimination (AUC). We will also compare its performance directly to existing definitions of screening eligibility including USPSTF criteria and the PLCOm2012 risk model.^14^ Sensitivity analyses will exclude late-stage cases with blood draw close to diagnosis.

##### Nodule Malignancy project

The goal of the Nodule Malignancy project is to identify biomarkers that can differentiate benign versus malignant pulmonary nodules, and the study design is based on the following considerations. First, to focus on the actionable time window while maximizing sample size, we included cases diagnosed within 5 years following blood draw. For lung cancers diagnosed at the baseline screen, the sample collected at baseline was included. This differs from post-diagnostic samples because all individuals participating in LDCT screening are without cancer diagnosis and mostly asymptomatic at baseline. Second, to maximize statistical power and ensure robust discovery results, we included 4 of the LDCT screening studies in the expanded targeted discovery phase (**Figure 1**). Third, the main comparison group is comprised of individuals with benign nodules who did not develop lung cancer, frequency matched on age at enrollment, age at the abnormal finding, age at blood collection, sex, and follow-up time. When multiple study participants with nodules were available as the matched benign nodule-control, we chose participants with higher estimated probability of nodule malignancy based on the Brock model to increase power for nodules with higher malignancy potential.^19^ To examine levels of proteins among nodule-free individuals in the screening-eligible population, we also included one control with no nodule findings per case, frequency matched on age at enrollment, age of blood collection, sex, and follow-up time.

###### Targeted discovery phase

The Nodule Malignancy project used a broad targeted discovery phase. We measured all available panels except the Cell Regulation panel, which did not show any robust associations with lung cancer in the Risk Biomarker project full discovery phase (**Table 2**). We included samples from the Pan-Canadian Early Detection of Lung Cancer Study (PanCan), UK Lung Cancer Pilot Screening Trial (UKLS), International Early Lung Cancer Action Program (IELCAP)-Toronto, and Pamplona-IELCAP (**Table 1;** further details in **Supplementary Table 1**). All samples within each LDCT study were randomly plated regardless of their cancer or nodule status to avoid batch effects by case status.

Within each study, protein measurements were standardized by z-transformation prior to pooled analysis. NPX values that did not pass QC were removed. We conducted multivariable logistic regression for each protein, adjusting for the Brock nodule malignancy score which includes age, sex, family history of lung cancer, emphysema, and nodule size, type, location, count, and spiculation (when available).^19^

To select protein markers for the INTEGRAL panel, we are using elastic net penalized regression^30^ and a random-forest-based feature selection approach^31^ to identify the combination of markers that best predicts nodule malignancy. We will also conduct analyses stratified by time to diagnosis. We will prioritize markers based on selection by either elastic net or random forest, consistency of results across studies, and association with lung cancer diagnosis within 1 year.

###### Validation phase

To evaluate the results obtained from the targeted discovery based on relative abundance, we will measure the INTEGRAL panel with absolute quantification in the same set of samples (PanCan, UKLS, IELCAP-Toronto, Pamplona-IELCAP), plus 1 independent study, the Pittsburgh Lung Screening Study (PLuSS). The model will be trained on the 4 original studies, and then evaluated in the PLuSS study. This enables evaluation of the data using absolute quantification of the protein markers (using the same set of studies), as well as external validation of the predictive accuracy (using the independent study).

### Harmonized databases created within the framework of the INTEGRAL Risk Biomarker and Nodule Malignancy projects

#### Risk Biomarker Project

One challenge for implementing risk-model-based eligibility for lung cancer screening is the unclear generalizability of risk prediction models in diverse worldwide populations.^13,14,32^ We therefore leveraged the infrastructure from the Risk Biomarker project and the Lung Cancer Cohort Consortium to develop a comprehensive study database for lung cancer incidence and mortality.

The cohorts contributing data on all participants to the LC3 harmonized database include most cohorts in the Risk Biomarker project and some additional cohorts. In total, 24 cohorts have contributed data on nearly 3 million participants (**Table 3**, descriptions in **Supplement**). The years of enrollment range from 1985 to 2010 and geographical regions include North America, Europe, Asia, and Australia. More than 69,000 lung cancer cases have been diagnosed during follow-up, including over 7,600 cases among people who never smoked.

**Table 3:** Description of the harmonized Lung Cancer Cohort Consortium database.

| Cohort | Location | Years of enrollment | Participants, N | Median follow-up (years)* | Female participants, % | Age at enrollment, median (min-max) | ----- Lung cancer cases, N (%) ----- |  |  |  |
| --- | --- | --- | --- | --- | --- | --- | --- | --- | --- | --- |
|  |  |  |  |  |  |  | Total** | Never smoking | Former smoking | Current smoking |
| AARP | USA | 1995-1996 | 565,645 | 15.5 | 40% | 62 (50-71) | 28,652 | 2,124 (8) | 15,272 (55) | 10,189 (37) |
| ATBC | Finland | 1985-1988 | 29,133 | 17.7 | 0% | 57 (49-70) | 3,959 | - | - | 3,959 (100) |
| CLUE | USA | 1989 | 30,461 | 29.1 | 57% | 48 (18-101) | 762 | 69 (9) | 271 (36) | 422 (55) |
| CPS-II | USA | 1992-1993 | 144,670 | 13.8 | 55% | 70 (47-90) | 3,745 | 446 (12) | 2,519 (67) | 778 (21) |
| CSDLH | Canada | 1992-1998 | 11,189 | 12.3 | 49% | 62 (23-100) | 367 | 65 (18) | 203 (56) | 93 (26) |
| EPIC | Europe | 1992-2000 | 518,112 | 14.9 | 71% | 51 (19-98) | 5,233 | 610 (12) | 1,468 (28) | 3,155 (60) |
| GCS | Iran | 2004-2008 | 50,032 | 13.0 | 58% | 52 (36-78) | 118 | 53 (45) | 4 (3) | 61 (52) |
| GS | UK | 2003-2009 | 106,761 | 9.6 | 100% | 47 (18-102) | 217 | 57 (29) | 87 (44) | 52 (27) |
| HPFS | USA | 1986 | 50,444 | 25.2 | 0% | 55 (32-81) | 1,295 | 164 (13) | 635 (51) | 444 (36) |
| HUNT | Norway | 1995-1997 | 78,941 | 16.9 | 53% | 48 (19-101) | 719 | 34(5) | 167 (24) | 504 (71) |
| MCCS | Australia | 1990-1994 | 41,473 | 23.1 | 59% | 55 (28-76) | 855 | 139 (16) | 377 (44) | 338 (40) |
| NHS | USA | 1976 | 120,617 | 39.9 | 100% | 43 (29-56) | 3,986 | 383 (10) | 489 (12) | 3,103 (78) |
| NYUWHS | USA | 1985-1991 | 14,266 | 30.0 | 100% | 50 (31-70) | 484 | 77 (18) | 166 (38) | 194 (44) |
| PHS | USA | 1982 | 26,338 | 11.7 | 0% | 65 (50-99) | 228 | 49 (21) | 127 (56) | 52 (23) |
| PLCO | USA | 1993-2001 | 154,884 | 11.9 | 50% | 63 (49-78) | 3,827 | 311 (8) | 1,821 (50) | 1,551 (42) |
| SCCS | USA | 2002-2009 | 84,429 | 11.2 | 60% | 52 (40-79) | 1,846 | 109 (6) | 369 (21) | 1,316 (73) |
| SCHS | Singapore | 1999-2003 | 50,962 | 13.5 | 57% | 63 (46-86) | 1,300 | 393 (30) | 267 (21) | 640 (49) |
| SCS | China | 1986-1989 | 18,069 | 25.3 | 0% | 56 (31-79) | 1,098 | 167 (15) | 69 (6) | 862 (79) |
| SMHS | China | 2002-2006 | 61,469 | 12.2 | 0% | 55 (40-75) | 1,164 | 173 (15) | 178 (15) | 813 (70) |
| SWHS | China | 1996-2000 | 79,940 | 18.1 | 100% | 50 (40-70) | 975 | 898 (92) | 12 (1) | 65 (7) |
| UKBB | UK | 2006-2010 | 502,105 | 12.1 | 54% | 57 (37-73) | 4,094 | 728 (18) | 1,764 (44) | 1,550 (38) |
| VITAL | USA | 2000-2002 | 77,118 | 10.0 | 52% | 62 (50-77) | 1,374 | 110 (8) | 782 (58) | 450 (34) |
| WHI | USA | 1993-1998 | 118,749 | 18.2 | 100% | 64 (49-83) | 2,389 | 415 (18) | 1,371 (58) | 574 (24) |
| WHS | USA | 1992-1995 | 39,852 | 24.1 | 100% | 55 (39-90) | 588 | 91 (15) | 200 (34) | 297 (51) |
| **Total** |  |  | **2,970,659** |  |  |  | **69,275** | **7,665 (11)** | **28,618 (42)** | **31,462 (47)** |

\*Follow-up time for lung cancer incidence. Mortality follow-up time may differ.
\*\*Cases with missing smoking status are included in the total, but not the stratified counts, so in some cases the stratified counts may not sum to the total.
Details on the eligibility criteria, data collection, and outcome ascertainment for each cohort are described in the **Supplement**. Time varying variables such as age were assessed as of the time of blood draw, or if blood was not collected, as of enrollment. Participants with a history of lung cancer prior to enrollment were excluded. For CSLDH,
the dataset provided is a case-cohort sample (see **Supplement**). For SCHS, the initial enrollment took place during 1993-1998, but the 1999-2003 follow-up visit was used as the baseline for the LC3 dataset (further information in **Supplement**). For WHI, the data include the observational study and the control arms of the Clinical Trials.
AARP: NIH-AARP Diet and Health Study; ATBC: Alpha-Tocopherol, Beta-Carotene Cancer Prevention Study; CLUE: Campaign Against Cancer and Heart Disease II; CPS-II: American Cancer Society Cancer Prevention Study-II Nutrition Cohort; CSDLH: Canadian Study of Diet, Lifestyle and Health; EPIC: European Prospective Investigation into Cancer and Nutrition; GCS: Golestan Cohort Study; GS: Generations Study; HPFS: Health Professionals Follow-up Study; HUNT2 & HUNT3: Trøndelag Health Study; MCCS: Melbourne Collaborative Cohort Study; NHS: Nurses' Health Study I and II; NYUWHS: New York University Women's Health Study; PHS: Physician's Health Study; PLCO: Prostate, Lung, Colorectal, and Ovarian Cancer Screening Trial; SCCS: Southern Community Cohort Study; SCHS: Singapore Chinese Health Study; SCS: Shanghai Cohort Study; SMHS: Shanghai Men's Health Study; UKBB: UK Biobank; VITAL: VITamins And Lifestyle Study; WHI: Women's Health Initiative; WHS: Women's Health Study.

Details on the eligibility criteria, data collection, and outcome ascertainment for each cohort are provided in the **Supplement** and the list of variables in **Table 4**. The variables were chosen to maximize our ability to calculate risk estimates for existing lung cancer prediction models.^33,34^ We applied a harmonization protocol aiming to minimize missing data and maintain consistent definitions while preserving data granularity. An initial analysis in the harmonized dataset compared the performance of lung cancer risk models in the United Kingdom.^35^

**Table 4:** Variables included in the harmonized databases for the Lung Cancer Cohort Consortium (Risk Biomarker project) and LDCT screening studies (Nodule Malignancy project)

| **Variables included in the harmonized Lung Cancer Cohort Consortium database (Risk Biomarker project)** |  |  |  |  |
| --- | --- | --- | --- | --- |
| **Demographic information** | **Follow-up and outcomes** | **Smoking** | **Exposures other than smoking** | **Personal health history** |
| • Age<br>• Sex<br>• Education<br>• Race/ethnicity<br>• Year of enrollment or blood draw<br>• State or region of residence (for USA cohorts) | • Follow-up time for lung cancer and death<br>• Lung cancer diagnosis with TNM stage and histology<br>• Vital status and cause of death, including lung cancer death | • Smoking status<br>• Years smoked<br>• Age at smoking initiation<br>• Age at smoking cessation<br>• Years since quitting<br>• Pack-years smoked<br>• Smoking intensity (cigarettes per day)<br>• Type of tobacco product<br>• Time to first cigarette | • Secondhand smoke exposure<br>• Asbestos exposure<br>• Indoor air pollution (e.g. cookstoves) | • Body mass index<br>• Family history of lung cancer<br>• Personal history of cancer<br>• COPD or emphysema<br>• Asthma<br>• Tuberculosis<br>• Daily cough<br>• Liver or kidney condition<br>• Diabetes<br>• Chronic bronchitis<br>• Hypertension<br>• Stroke<br>• Heart attack or heart disease |
| **Variables included in the harmonized LDCT screening study database (Nodule Malignancy project)** |  |  |  |  |
| **Demographic information** | **Follow-up and outcomes** | **Smoking** | **Nodule characteristics** | **Personal health history** |
| • Age<br>• Sex<br>• Education<br>• Race/ethnicity<br>• Country | • Follow-up time for lung cancer and death<br>• Lung cancer diagnosis with TNM stage and histology<br>• Vital status and cause of death, including lung cancer death | • Smoking status<br>• Duration of smoking<br>• Age at smoking initiation<br>• Age at smoking cessation<br>• Years since quitting<br>• Pack-years smoked<br>• Smoking intensity (cigarettes per day) | • Screening round<br>• Date of screening<br>• Nodule location<br>• Nodule size<br>• Attenuation<br>• Nodule count<br>• Semantic features (spiculation, margin, calcification)<br>• Malignant status | • Body mass index<br>• Family history of lung cancer<br>• Personal history of cancer<br>• COPD<br>• Spirometry measures<br>• Asthma<br>• Chronic bronchitis |

We have defined a priority to facilitate sharing of the LC3 harmonized database, with the vision that it will serve as a resource for future research on lung cancer. We are currently establishing a legal and technical infrastructure that will allow investigators outside of the LC3 consortium to request permission to remotely access and analyze the data in a secure computing environment. Available data will include the variables listed in **Table 4**, the metabolomics biomarkers measured in the first project of the LC3,^36^ and eventually the proteomics biomarkers following the publication of the validation phase of the project.

#### Nodule Malignancy Project

For the Nodule Malignancy project, data from 6 LDCT screening studies were harmonized within the framework of ILCCO. In addition to the 5 LDCT screening studies described above, the National Lung Screening Trial (NLST) is also participating in the Nodule Malignancy project for quantitative imaging analysis. The design of each CT screening program including eligibility and recruitment framework is described in the **Supplement**.

For quality control, the data from all studies were systematically checked for missing values, outliers, inadmissible values, aberrant distributions, and internal inconsistencies. All procedures were recorded for each study and a central data dictionary is maintained throughout the process. A total of 2,088 cases and 42,940 screened individuals from the 6 LDCT screening studies are included in the harmonized database of screening studies (**Supplementary Table 2**). The variables that are compatible across the screening studies are shown in **Table 4**.

### Perspectives

With the advent of LDCT screening, the potential to substantially reduce lung cancer mortality has vastly expanded, and so has the domain of potential research questions. The current work of the INTEGRAL program aims to address two specific ways in which biomarkers might contribute; namely, to improve the selection of individuals for screening, and to better distinguish between malignant and benign nodules on LDCT images. At the completion of our current work, we anticipate that we will have developed a fit-for-purpose biomarker panel that can be applied in both settings. For pre-screening risk assessment, we will deliver an integrated risk prediction model including the biomarkers on the panel and results of a comprehensive independent validation study of its performance. For nodule discrimination, we will establish an integrated nodule probability model including quantitative radiological features and biomarkers.

If these steps are successful, important work will remain to implement the INTEGRAL panel in clinical practice. Specific considerations related to biomarker implementation have been outlined.^37^ We plan to assess whether repeated measurements of the panel could improve our ability to predict lung cancer risk. Implementation studies will be needed to determine the feasibility of this approach in practice. The design of future evaluations will require careful consideration, as we consider it infeasible to evaluate the incremental improvement in performance offered by biomarkers in the setting of a randomized trial. Finally, another future goal might be to identify predictors of lung cancer among people are light smokers or never smoked, which could be used to evaluate patients with symptoms potentially suspicious for lung cancer.

It is important to note that many other tools exist or are being developed to refine risk estimation for lung cancer, including both biomarkers and risk prediction models. Another important future direction will be to directly compare the performance of these tools or, where feasible and cost-effective, to integrate them. Comparisons should be made in the same set of samples so that discrimination metrics can be directly compared.

The INTEGRAL biomarker program represents an ambitious initiative to develop a flexible biomarker tool to improve early lung cancer detection via LDCT screening. With a focus on protein biomarkers, the program spans discovery, panel development, model training and validation – all whilst remaining in an observational framework. The forthcoming results from the validation phase of INTEGRAL will provide a definitive benchmark on the potential for circulating protein biomarkers to improve early detection of lung cancer – and most importantly – whether it is justified to introduce them in a screening scenario to inform who should be screened and how to manage nodules.

## Data Availability

Data in the Lung Cancer Cohort Consortium harmonized database will soon be available by secure remote access following submission and approval of a research proposal. For more information, please contact the first author.

