## Supplemental Material for "Design and methodological considerations for biomarker discovery and validation in the Integrative Analysis of Lung Cancer Etiology and Risk (INTEGRAL) Program"

#### SUPPLEMENT

**Supplementary Table 1: Further description of lung cancer cases participating in the development and validation of the INTEGRAL protein panel for lung cancer early detection and nodule malignancy *(extension of Table 1)***

| Study component | Lung cancers by histological* type | | | Lung cancers by sex | | Lung cancers by lead time prior to diagnosis | | |
|---|---|---|---|---|---|---|---|---|
| | Adenocarcinoma | Squamous cell carcinoma | Small cell carcinoma | Male | Female | <1 year | 1-2 years | 2-3 years |
| **Risk Biomarker: Full discovery** | | | | | | | | |
| European Prospective Investigation into Cancer and Nutrition (EPIC) | 58 (31%) | 34 (18%) | 35 (19%) | 127 (68%) | 61 (32%) | 55(29%) | 55 (29%) | 78 (41%) |
| Northern Sweden Health and Disease Study (NSHDS) | 23 (36%) | 13 (20%) | 6 (9%) | 32 (50%) | 32 (50%) | 15 (23%) | 21 (33%) | 28 (44%) |
| **Total** | **81 (32%)** | **47 (19%)** | **41 (16%)** | **159 (63%)** | **93 (37%)** | **70 (28%)** | **76 (30%)** | **106 (42%)** |
| **Risk Biomarker: Targeted discovery*** | | | | | | | | |
| Cancer Prevention Study II (CPS-II) | 38 (33%) | 27 (23%) | 16 (14%) | 73 (63%) | 42 (37%) | 30 (26%) | 45 (39%) | 40 (35%) |
| Nord-Trøndelag Health Study (HUNT) | 54 (33%) | 34 (21%) | 26 (16%) | 101 (62%) | 63 (38%) | 58 (35%) | 48 (29%) | 58 (35%) |
| Melbourne Collaborative Cohort Study (MCCS)** | 49 (45%) | 19 (18%) | 21 (19%) | 75 (70%) | 33 (30%) | 41 (38%) | 37 (34%) | 30 (28%) |
| Singapore Chinese Health Study (SCHS) | 24 (26%) | 23 (25%) | 14 (15%) | 82 (89%) | 10 (11%) | 28 (30%) | 29 (32%) | 35 (38%) |
| **Total** | **165 (34%)** | **103 (22%)** | **77 (16%)** | **331 (69%)** | **148 (31%)** | **157 (33%)** | **159 (33%)** | **163 (34%)** |
| **Risk Biomarker: Validation – training set*** | | | | | | | | |
| Campaign Against Cancer and Heart Disease (CLUE) | 19 (32%) | 7 (12%) | 9 (15%) | 34 (57%) | 26 (43%) | 20 (33%) | 22 (37%) | 18 (30%) |
| Cancer Prevention Study II (CPS-II) | 38 (33%) | 27 (23%) | 16 (14%) | 73 (63%) | 42 (37%) | 30 (26%) | 45 (39%) | 40 (35%) |
| Nord-Trøndelag Health Study (HUNT) | 54 (33%) | 34 (21%) | 26 (16%) | 101 (62%) | 63 (38%) | 58 (35%) | 48 (29%) | 58 (35%) |
| Melbourne Collaborative Cohort Study (MCCS)** | 49 (45%) | 19 (18%) | 21 (19%) | 75 (70%) | 33 (30%) | 41 (38%) | 37 (34%) | 30 (28%) |
| Physicians' Health Study (PHS) | Missing | Missing | Missing | 29 (100%) | 0 (0.0%) | 9 (31%) | 12 (41%) | 8 (28%) |
| Singapore Chinese Health Study (SCHS) | 24 (26%) | 23 (25%) | 14 (15%) | 82 (89%) | 10 (11%) | 28 (30%) | 29 (32%) | 35 (38%) |
| Women's Health Initiative (WHI)** | 197 (44%) | 67 (15%) | 49 (11%) | 0 (0.0%) | 445 (100%) | 116 (26%) | 166 (37%) | 163 (37%) |
| **Total** | **381 (38%)** | **177 (17%)** | **135 (13%)** | **394 (39%)** | **619 (61%)** | **302 (30%)** | **359 (35%)** | **352 (35%)** |
| **Risk Biomarker: Validation – testing set** | | | | | | | | |

| | | | | | | | | |
|---|---|---|---|---|---|---|---|---|
| Alpha-Tocopherol, Beta-Carotene Cancer Prevention Study (ATBC) | 47 (14%) | 145 (44%) | 84 (26%) | 327 (100%) | - | 68 (21%) | 113 (35%) | 146 (45%) |
| Golestan Cohort Study (GCS) | Missing | Missing | Missing | 13 (93%) | 1 (7%) | 5 (36%) | 8 (57%) | 1 (7%) |
| New York University Women's Health Study (NYUWHS) | 9 (47%) | 2 (11%) | 2 (11%) | - | 19 (100%) | 4 (21%) | 10 (53%) | 5 (26%) |
| Shanghai Cohort Study (SCS) | 11 (20%) | 21 (38%) | 3 (5%) | 56 (100%) | - | 24 (43%) | 16 (29%) | 16 (29%) |
| Southern Community Cohort Study (SCCS) | 38 (27%) | 36 (25%) | 21 (15%) | 81 (57%) | 62 (43%) | 36 (25%) | 50 (35%) | 56 (40%) |
| Shanghai Men's Health Study (SMHS) | 38 (42%) | 33 (36%) | 9 (10%) | 91 (100%) | - | 30 (33%) | 29 (32%) | 32 (35%) |
| Women's Health Study (WHS) | 14 (42%) | 4 (12%) | 5 (15%) | - | 33 (100%) | 14 (42%) | 11 (33%) | 8 (24%) |
| **Total** | **157 (23%)** | **241 (36%)** | **124 (19%)** | **568 (83%)** | **115 (17%)** | **181 (26%)** | **237 (35%)** | **264 (39%)** |

| | **Adenocarcinoma** | **Squamous cell carcinoma** | **Small cell carcinoma** | **Male** | **Female** | **≤1 year** | **>1 to 3 years** | **>3 to 5 years** |
|---|---|---|---|---|---|---|---|---|
| **Nodule Malignancy: Targeted discovery & validation** | | | | | | | | |
| Pan-Canadian Early Detection of Lung Cancer Study (PanCan) | 97 (57%) | 21 (12%) | 11 (7%) | 83 (49%) | 86 (51%) | 65 (39%) | 79 (47%) | 25 (15%) |
| The UK Lung Cancer Pilot Screening Trial (UKLS) | 48 (48%) | 26 (26%) | 10 (10%) | 80 (79%) | 21 (21%) | 49 (49%) | 27 (27%) | 25 (25%) |
| The International Early Lung Cancer Action Program (IELCAP-Toronto) | 46 (58%) | 7 (9%) | 2 (3%) | 23 (29%) | 56 (71%) | 36 (46%) | 29 (37%) | 11 (14%) |
| The International Early Lung Cancer Action Program (IELCAP-Spain) | 36 (47%) | 15 (20%) | 5 (7%) | 63 (83%) | 13 (17%) | 36 (47%) | 23 (30%) | 16 (21%) |
| Validation: The Pittsburgh Lung Screening Study (PLuSS) | 97 (39%) | 57 (23%) | 33 (13%) | 151 (60%) | 99 (40%) | 138 (55%) | 63 (25%) | 49 (20%) |
| **Total** | **324 (48%)** | **126 (19%)** | **61 (9%)** | **400 (59%)** | **275 (41%)** | **324 (48%)** | **221 (33%)** | **126 (19%)** |

*Cohorts in the Risk Biomarker targeted discovery phase are also included in the validation phase training set and are listed twice in the table.
**In MCCS and WHI, participants were sampled separately at two different blood draws. For the stratified selection of subcohort representatives, WHI included a stratification by study arm (observational study or the non-intervention arm of the clinical trial).
INTEGRAL, the Integrative Analysis of Lung Cancer Etiology and Risk program
IELCAP, the International Early Lung Cancer Action Program
For the Risk Biomarker project, lung cancer cases with other and missing histological type are excluded from the table. These numbers are, respectively in each cohort, 69 and 1 in EPIC, 7 and 15 in NSHDS, 45 and 0 in CPS-II, 52 and 6 in HUNT, 27 and 0 in MCCS, 21 and 11 in SCHS, 20 and 12 in CLUE, 0 and 29 in PHS, 179 and 1 in WHI, 53 and 9 in ATBC, 0 and 14 in GCS, 10 and 0 in NYUWHS, 6 and 17 in SCS, 50 and 0 in SCCS, 16 and 0 in SMHS, 6 and 4 in WHS.

**Supplementary Table 2: Description of the harmonized database of LDCT screening studies from the INTEGRAL Nodule Malignancy project**

| Cohort | Location | Screening period | Participants, N | Lung cancer cases, N | Age | Participants by sex, N (%) | | Participants by smoking status, N (%) | | |
|---|---|---|---|---|---|---|---|---|---|---|
| | | | | | Median (min-max) | Male | Female | Never smoking | Former smoking | Current smoking |
| **NLST** | US | 2002-2007 | 26722 | 1071 | 60 (55-74) | 15770 (59) | 10952 (41) | 0 (0) | 13860 (52) | 12862 (48) |
| **PLuSS** | US | 2002-2016 | 3642 | 391 | 58 (50-80) | 1872 (51) | 1770 (49) | 0 (0) | 1088 (30) | 2554 (70) |
| **PanCan** | Canada | 2008-2014 | 2537 | 178 | 62 (50-76) | 1401 (55) | 1136 (45) | 0 (0) | 945 (37) | 1592 (63) |
| **IELCAP-Toronto** | Canada | 2003-2019 | 4161 | 206 | 60 (47-83) | 1869 (45) | 2292 (55) | 9 (0) | 2691 (65) | 1424 (35) |
| **UKLS** | England | 2011-2013 | 2053 | 145 | 68 (55-76) | 1551 (76) | 502 (24) | 5 (0) | 1288 (63) | 760 (37) |
| **Pamplona-IELCAP** | Spain | 2001-2020 | 3825 | 97 | 55 (31-87) | 2728 (71) | 1094 (29) | 0 (0) | 1444 (38) | 2378 (62) |
| **Total** | | | **42,940** | **2088** | | **25191 (59)** | **17746 (41)** | **14 (0)** | **21316 (50)** | **21570 (50)** |

IELCAP: International Early Lung Cancer Action Program; PanCan: Pan-Canadian Early Detection of Lung Cancer Study; PLuSS: Pittsburgh Lung Screening Study; NLST: National Lung Screening Trial; UKLS: United Kingdom Lung Cancer Screening Trial

### Descriptions of prospective cohort studies participating in the INTEGRAL Risk Biomarker project and/or in the Lung Cancer Cohort Consortium harmonized database

**AARP: NIH-AARP Diet and Health Study**

The NIH-AARP Diet and Health Study began in 1995 to study the relationship between diet and health.[1] A total of 566,398 participants were recruited from among the members of the American Association of Retired Persons aged 50-71 years who lived in six US states (California, Florida, Pennsylvania, New Jersey, Carolina and Louisiana) or in two urban areas (Atlanta, Georgia and Detroit, Michigan). The baseline questionnaire assessed information on lifestyle, diet, and medical history. Participants were not eligible if they reported a colorectal, breast, or prostate cancer, or a renal disease. Cancer cases are identified through linkage to cancer registries and vital status is recorded through the National Death Index.

**ATBC: Alpha-Tocopherol, Beta-Carotene Cancer Prevention Study**

The ATBC randomized trial enrolled 29,133 male participants from 1985 to 1988.[2] Men residing within the study region (Southwest Finland) and aged from 50 to 69 were mailed a questionnaire about their smoking habits and willingness to participate. Eligibility required active smoking of at least 5 cigarettes per day, with no prior cancer (except nonmelanoma skin cancer), severe angina on exertion, chronic renal insufficiency, liver cirrhosis, chronic alcoholism, nor any other medical problems that might limit participation for 6 years, such as psychiatric disorder or physical disability. Participants were excluded if they were receiving anticoagulant therapy or using supplements containing vitamin E (>20 mg/d), vitamin A (>20000 IU/d = 4000 retinol equivalents), or beta-carotene (>6 mg/d). Enrolled participants were randomized to one of four treatment groups: alpha-tocopherol alone, alpha-tocopherol and beta-carotene, beta-carotene, alone, or placebo with a 1:1:1:1 randomization ratio. At baseline, participants completed questionnaires regarding general risk factors, medical history, smoking habits, and dietary intake. Height, weight, heart rate, and blood pressure were measured by trained nurses and fasting serum samples were collected and stored at -70$^{o}$C. Supplementation continued for 5-8 years (median 6.1 years) until death or the end of the trial (April 30, 1993). Identification of incident cancers was primarily via the Finnish Cancer Registry, which provides almost 100% case ascertainment nationwide. Participants were also asked to contact their field center if diagnosed with cancer. Post-intervention follow-up has continued through the Finnish Cancer Registry and other national registries. For the LC3, we included participants in all four study arms.

**CLUE: Campaign Against Cancer and Heart Disease II**

The Campaign Against Cancer and Heart Disease is a community-based cohort that enrolled almost 33,000 volunteer participants aged 2-98 years in Washington County, Maryland, USA in 1989.[3] At enrollment, participants provided information including age, race, sex, marital status, time since last menstrual period, smoking history, blood pressure, medication use, hormone use, height and weight currently and at age 21, and medical history. Follow up was conducted via questionnaires administered in 1996, 1998, 2000, 2003, and 2007. Cancer diagnoses were ascertained through regular linkage to the Washington County hospital records and state cancer registry. Deaths were identified via hospital records, Maryland Vital Statistics, National Death Index, next of kin, and obituaries. At enrollment, a 20 mL blood specimen was drawn and stored at -70°C.

**CPS-II: American Cancer Society Cancer Prevention Study-II Nutrition Cohort**

CPS-II was a prospective study of cancer initiated in 1982 in all 50 US states.[4] It included 1.2 million American men and women. In 1992, the Nutrition study was established as a subgroup of the larger CPS-II with the main objectives of obtaining detailed information on dietary and other exposures, identifying incident cases of cancer and deaths, and evaluating dietary and other risk factors for cancer in a large prospective study. Between 1992 and 1993, participants completed a 10-page self-administered questionnaire that included demographic, medical history, dietary and lifestyle information. The cohort ultimately included 86,406 men and 97,788 women ages 50–74 years in 21 states with population-based state cancer registries reported to ascertain at least 90% of incident cancer cases. Between June 1998 and June 2001, blood samples were collected from a subset of CPS-II Nutrition Cohort participants (21,965 women and 17,411 men) and stored at -130°C. Incident cancers were identified either by self-report on a follow-up questionnaire (sent every 2 years via email), linkage with state registries, or death certificates. Mortality follow-up is done by linkage with the National Death Index.

**CSDLH: Canadian Study of Diet, Lifestyle and Health**
The CSDLH is a prospective cohort that began in 1992, with the aim to study the link between diet, lifestyle factors, molecular markers and cancer risk in Canada.[5] The 73,909 participants were mainly recruited through the alumni associations of three universities (Toronto, Alberta and Western Ontario), while a small number of participants was recruited through the Canadian Cancer Society (n=5687). Eligibility required age 18 years or older and there were no other exclusion criteria. At enrollment, participants completed a food frequency questionnaire and a lifestyle questionnaire that sought information on lifestyle factors, medical history, physical measures and demographics. Cancer cases and vital status are ascertained by linkage to Canadian national and provincial registries. CSDLH contributed a case-cohort sample (cases and an age-stratified random subcohort sample) to the LC3 in place of the full cohort database.

**EPIC: European Prospective Investigation into Cancer and Nutrition**
The EPIC study is a large prospective cohort with more than 521,330 participants enrolled at 23 centers in 10 western European countries between 1992 and 2000.[6–8] Eligibility in each center was based on age, gender, and geographic or administrative boundaries.[7] The age range was generally from 35 to 70 years and participants were invited to participate either by mail or in person. Detailed information on diet, lifestyle characteristics, anthropometric measurements, and medical history was collected at recruitment. Biological samples including plasma, serum, leukocytes, and erythrocytes were also collected from approximately 388,000 individuals and stored in liquid nitrogen at -196°C , except in Umea, Sweden, where samples were stored at -80°C and in Denmark, where samples were stored in liquid nitrogen vapor at -150°C. Follow-up to identify cancer cases is based on population cancer registries in seven of the participating countries (Denmark, Italy, The Netherlands, Norway, Spain, Sweden and the UK) and on a combination of methods including health insurance records, cancer and pathology registries, and active follow-up through study subjects and their next-of-kin in three countries (France, Germany and Greece). In parallel, data on total and cause-specific mortality are collected at the EPIC study centers through mortality registries or active follow-up and death-record collection.

**GCS: Golestan Cohort Study**
The Golestan Cohort Study is a prospective cohort aiming to study risk factors for chronic diseases, with an emphasis on oesophageal cancer, in the Golestan province in northeastern Iran.[9] Participants were selected randomly by systemic clustering, using household numbers, and were then contacted and invited by trained staff to participate in the study. Participation rates were 84% for women and 70% for men. From 2004 to 2008, the study enrolled 50,045 participants aged 36 to 80. Eligibility required no prior history of gastrointestinal cancers and permanent residence in the province of Golestan. Baseline data

was collected by trained physicians and nutritionist via interviews. Participants provided hair, urine, nail and blood specimens. 10 mL of blood was collected from participants and stored at -80°C. Participants have been actively followed every 12 months and were also instructed to contact the study in case of important events such as hospitalizations or development of major diseases. Clinical outcomes are assessed by internal physicians who are independent from the study.

**GS: Generations Study**
The Generations Study enrolled over 113,000 women from the general population in the United Kingdom beginning in 2003, to study the causes of breast cancer.[10] Eligibility required age 16 years or older. Women who joined the study were asked to nominate a female acquaintance who was then contacted to join the study. The recruitment process involved a mailed questionnaire to the involved women and a blood draw, and active follow-up is done approximately every 3 years to update the risk factor information and collect follow-up information on breast cancer. Cancers cases and deaths are identified through the linkage to the National Health Service Central Registers.

**HPFS: Health Professionals Follow-up Study**
The Health Professionals Follow-up Study began in 1986 to evaluate the impact of nutritional factors on cancer, heart disease, and other cardiovascular diseases among men.[11] HPFS ultimately enrolled 51,529 male health professionals aged 40-75. The questionnaire at recruitment assessed detailed dietary, lifestyle, physical activity, and medical information. In addition, between 1993 and 1995, blood samples were collected for a subset of the population (18,225 participants) who were free of cancer, diabetes and heart diseases. Cancer cases and deaths were identified through linkage with diseases files, cancer files and mortality files.

**HUNT2 & HUNT3: Trøndelag Health Study**
The HUNT study is a large population-based cohort covering 125,000 Norwegian participants from three sub-cohorts: HUNT1 (enrollment 1984-86), HUNT2 (1995-97) and HUNT3 (2006-08).[12] For each subcohort, every citizen of Nord-Trøndelag County aged 20 years or older (or turning 20 years during the year of survey) was invited during the recruitment period. The study was primarily set up to address arterial hypertension, diabetes, screening of tuberculosis, and quality of life; however, the focus has expanded to include additional diseases. HUNT2 constituted both a new cross-sectional survey and a follow-up of HUNT1. The scientific programme was extended to include several large public health issues in accordance with current national health priorities, including cardiovascular disease, diabetes, obstructive lung disease, osteoporosis, headache, mental health, chronic musculoskeletal pain and urinary incontinence. In addition to questionnaires, interviews and clinical examinations, the participants contributed with blood samples stored at -80°C. A total of 65,237 individuals participated in HUNT2 (69.5% of those invited). In HUNT3, 93,860 residents were invited and 50,807 participated (54.1% participation rate). The scientific programme of HUNT3 included several public health issues as in HUNT2, but also included topics like cultural participation and religious affiliation. Optimal handling and storage of blood and urine samples were given high priority, as part of the establishment of a new modern biobank. The unique 11-digit identification number of every Norwegian citizen enabled linkage between the collected information and several local, regional or national registers. Incident lung cancer cases are ascertained based on the linkage of data between HUNT and the Cancer Registry of Norway while vital status (alive, emigrated and dead) and cause-specific deaths were collected by linkage to the Cause of Death Register.

**MCCS: Melbourne Collaborative Cohort Study**
MCCS is a prospective population cohort set up to investigate the role of diet and lifestyle in causing cancer and other non-communicable diseases and to investigate possible interactions between these exposures and common genetic variants.[13] Between 1990 and 1994, 41,513 residents of the Melbourne metropolitan area (24,469 women and 17,044 men) aged between 40 and 69 were recruited. All participants are of White European origin and about 24% of the cohort is Southern European migrants to Australia who were deliberately over-sampled to extend the range of lifestyle exposures and to increase genetic variation. At baseline, participants completed questionnaires that measured demographic characteristics and lifestyle factors and most (N=41,133) also donated a blood sample. Incident diagnosis of cancer and deaths were identified by record linkage to the Victorian Cancer Registry (VCR) and the Victorian Registry of Births, Deaths and Marriages. Both registries are considered complete, with notification to the VCR of all cancers diagnosed in Victoria mandated by legislation since 1981. There were two waves of active follow-up: the first wave of follow-up occurred between 1995 and 1998, while the second wave occurred between 2003 and 2007. For our biomarker study, we analyzed samples collected at baseline and from the second active follow-up visit. In this second follow-up, a total of 28,240 (68%) of the original participants, 17,138 women and 11, 102 men, attended clinics organized at the Cancer Council Victoria premises near the Melbourne CBD for face-to face interviews and physical measurements; 3007 had died before they could be contacted. Those who attended this visit were younger and more likely to be born in an English-speaking country and had higher socioeconomic position than those who did not.

**NHS: Nurses' Health Study I and II**
The NHS is a large prospective cohort to evaluate the risk factors for major chronic diseases in women.[14] Recruitment began in 1976 and the study includes more than 121,700 married registered nurses from 11 states in the US. Eligibility required age 30-55 and that participants be married. Potential participants were identified by nursing boards who agree to share their member's contact information with NHS. The baseline questionnaire included information on contraceptive use, smoking, cancer, and heart diseases. Self-reported cancers were confirmed by a review of medical records. An active follow-up is done every two years with heath related topics, diet, and disease questions. Disease diagnosis reports and cause of death are confirmed by several methods including medical record reviews, supplemental questionnaires, and in some cases self-report. For cancers, following permission from the participant, the participant's doctor or hospital is contacted to request medical records and pathology reports. Blood samples were collected from 33,000 participants in 1989-1990 and stored between -130°C and -196°C. Death information is obtained by the linkage to the US National Death Index.

**NSHDS: Northern Sweden Health and Disease Study**
NSHDS is an ongoing prospective cohort and intervention study intended to promote health in the population of Västerbotten County in northern Sweden.[15] Study participants were recruited to the NSHDS in the context of the Västerbotten Intervention Project (VIP), which was initiated in 1985 to advocate a healthy diet and lifestyle.[16] In 1985, when the VIP began, all residents in Västerbotten County were invited to participate by attending a health check-up at 40, 50, and 60 years of age. At the health check-up, participants completed a self-administered questionnaire covering various factors such as education, smoking habits, physical activity, and diet. In addition, height and weight were measured and participants were asked to donate a fasting blood sample. Over 135,000 Västerbotten County residents participated in the health survey. At regular intervals the cohort is scanned for incident myocardial infarctions (MI) and stroke using the Northern Sweden MONICA registry and for cancer using the regional cancer registry. The same procedure has also started for other

registries, e.g. diabetes, osteoporosis, and Alzheimer's disease. For the biomarker studies, lung cancer cases were defined using the International Classification of Diseases for Oncology, Second Edition (ICD-O-2), and included all primary malignant cancers coded as C34.0-C34.9 with pre-diagnostic blood samples.

**NYUWHS: New York University Women's Health Study**
NYUWHS is a prospective study started in 1985 to study the causes of cancer and other chronic diseases in women.[17,18] The study enrolled 14,274 participants who donated a blood sample and completed a questionnaire about health history, diet, exercise, lifestyle habits and reproductive history. Blood samples were subsequently stored at -80°C. The study enrolled women aged 35-65 at the Guttman Breast Diagnostic Institute, a breast cancer screening center in New York City. Women who had been pregnant 6 months before the visit or taken hormonal medications were excluded. Cancer cases were reported through active follow-up every 2-4 years and confirmed by linkage with cancer registries in New York. Deaths were identified by the linkage to the US National Death Index.

**PHS: Physician's Health Study**
The Physician's Health Study (PHS) is a randomized trial started in 1982 to evaluate aspirin and beta carotene for the prevention of cardiovascular disease and cancer.[19] A second study was launched in 1997 to study other supplements including vitamin E, vitamin C, and multivitamins for primary prevention of cardiovascular disease, cancer, age-related eye disease, and cognitive decline. Potential participants were identified as male physicians, aged 40-84 years, who lived in the US and were registered with the American Medical Association. Physicians with an history of myocardial infarction, stroke, transient ischemic attack, cancer (except non-melanoma skin cancer), renal or liver disease, peptic ulcer, gout or contraindication to use aspirin or beta-carotene were excluded. The study finally enrolled 22,071 participants, who were randomly assigned to take aspirin and/or beta-carotene and/or placebo. The baseline questionnaire included information on lifestyle, medical history, and diet. During the first four months, participants were asked to donate a blood sample, and almost 15,000 blood samples were collected and frozen at -82°C. A follow-up questionnaire has been done at 6 months, 12 months, and annually thereafter. Cancer cases reported in questionnaires were confirmed by a subsequent review of medical records. Deaths were confirmed by hospital records and death certificates. Our study includes all randomization arms of the PHS.

**PLCO: Prostate, Lung, Colorectal, and Ovarian Cancer Screening Trial**
The PLCO trial is a randomized controlled trial evaluating the efficacy of screening for prostate, lung, colorectal, and ovarian cancers in reducing cancer mortality compared to usual healthcare.[20] PLCO enrolled 155,000 men and women aged 55-74 during 1993 to 2001 at 10 screening centers in the USA. Exclusion criteria included a history of prostate, lung, colorectal, or ovarian cancer, or surgical removal of the entire prostate, entire colon, or one lung. Enrolled participants were randomized 1:1 to screening programs for the four cancers or to usual care. At entry, participants completed a baseline questionnaire containing information such as demographics and medical history. An annual study update (ASU) was administered to the participants. Deaths occurring during the trial were mainly identified by ASUs or reports from relatives, friends, or physicians of participants, and followed by death certificate request and periodic linkage to the National Death Index (NDI). Cancers were primarily identified via ASUs or positive screening tests, and pathologically-confirmed through medical record review,[21] with later occurring cancers additionally identified by linkage to state tumour registries. Biological samples were collected from participants in the intervention arm at baseline and each of the next 5 years and stored at -157°C. Over the course of the PLCO trial, over 2.9 million biospecimens were collected.

**SCCS: Southern Community Cohort Study**
The Southern Community Cohort Study (SCCS) is a prospective cohort that was established to address many unresolved questions about the root causes of cancer health disparities.[22] From 2002-2009, the study enrolled approximately 85,000 adults aged 40-79 years from the southeastern states in the USA with the highest representation of African Americans (Alabama, Arkansas, Florida, Georgia, Kentucky, Louisiana, Mississippi, North Carolina, South Carolina, Tennessee, Virginia, and West Virginia.). Recruitment occurred mainly at Community Health Centers (CHCs), institutions providing basic health services primarily to the medically uninsured, so the cohort includes many adults of lower income and educational status. Each study participant enrolled at CHCs completed a detailed baseline in-person interview administered by trained personnel to collect information participant characteristics, behaviors, and health status. Nearly 90% of participants provided a biologic specimen (approximately 45% a blood sample, stored at -80°C, and 45% buccal cells). Follow-up for mortality is conducted by linkage to the National Death Index and Social Security mortality files. Follow-up for cancer incidence is conducted via linkage to state cancer registries. Participants also received follow-up questionnaires by mail about every 5 years.

**SCHS: Singapore Chinese Health Study**
SCHS is a population-based prospective cohort with a primary goal to elucidate the role of diet and its interaction with genetic factors in cancer development and other chronic diseases.[23] From 1993 to 1998, the study enrolled 63,257 men and women aged 45 to 74 years living in Singapore. At enrollment, each participating subject was interviewed in person at home by a trained interviewer using a structured questionnaire. The questionnaire collected information on demographics, body weight and height, history of cigarette smoking and use of other tobacco products, usual dietary intake habits, history of occupational exposures, exposure to incense as a measure of indoor air pollution, physical activity, sleep quantity and quality, menstrual and reproductive history (women only), medical history, and family history of cancer. New occurrences of cancers and deaths were regularly ascertained through linkage to the Singapore Cancer Registry and Singapore National Birth and Death Registry. During 1999 to 2003, a follow-up survey was conducted, and immediately afterwards a biospecimen collection was done for all participants who agreed to donate (including blood, urine, and/or mouthwash). During the study, 32,235 participants provided at least one biological sample (blood, urine, and/or mouthwash). In total 28,346 blood samples were collected. Multiple vials of each blood component, buccal cells, and urine samples were stored at -80°C immediately after processing. For our LC3 dataset, to facilitate the biomarker studies, the 1999-2003 follow-up visit was used as the baseline, and all time-varying variables were aligned to this baseline.

**SCS: Shanghai Cohort Study**
The SCS is a prospective residential cohort of men in Shanghai, China.[24] It enrolled 18,244 men aged 45-64 years during 1986-1989 who resided within geographically defined neighborhoods in the city of Shanghai. The primary goal of this study is to investigate the role of environmental and diet-related exposures quantified by biological markers in the causation of cancer. At the time of recruitment, each subject was interviewed in person by a trained nurse interviewer using a structured questionnaire to collect information on demographics, education, body weight and height, type of occupation, history of smoking of cigarettes and other tobacco products, history of alcohol use, dietary intake, and medical history. At recruitment, blood and a single void urine specimen were collected from all participants. Multiple vials of serum and urine samples per subject were created and stored at -72°C or below. New occurrences of cancer and death are regularly ascertained through

linkage to the Shanghai Cancer Registry and Shanghai Vital Statistics Units. In addition, follow-up interviews are performed by trained interviewers in person to all surviving subjects, or if the subject is deceased, with his next of kin. Follow-up interviews were done annually from 1990 to 2012, and biannually from 2013.

**SMHS: Shanghai Men's Health Study**
The SMHS is a population-based cohort study whose primary goal was to investigate contribution of lifestyle, environmental factors and genetic susceptibility to cancer and other non-communicable diseases.[25] It launched in 2002 in urban Shanghai and enrolled almost 62,000 men aged 40 to 74 years. At enrollment, in-person interviews were conducted to collect information on demographics, medical history, personal and dietary habits, physical activity, family history of cancer, occupational history, and weight and height history during adolescence and early adult years. A 10 mL blood sample was collected from 46,244 study participants who agreed to blood draw. All samples were processed within 6 hours and stored at -75°C. New occurrences of cancer are identified via regular searches of the Shanghai Cancer Registry. Follow-up for mortality is conducted via the Shanghai Vital Statistics Registry.

**SWHS: Shanghai Women's Health Study**
The SWHS is a large cohort study launched in 1996 in Shanghai for the purpose of studying the occurrence and etiology of cancer in Asian women.[26] From 1996 to 2000 the SWHS recruited more than 74,942 Chinese women aged 40-70. Participants were identified using a roster of all women aged 40-70 years obtained from the residency offices in the study community. All study participants completed a baseline questionnaire and conducted interviews with trained healthcare professionals to provide detailed data on lifestyle and dietary habits as well as body measurements. Consenting participants (about 88% of all participants) also provided urine, blood, and exfoliated buccal cells samples which are stored long term at -70°C. Follow-up information on study participants is gathered via biennial in-person recontact and regular linkage to cancer and vital statistics registries in Shanghai.

**UKBB: UK Biobank**
UK Biobank is a large, open-access, ongoing prospective cohort that enrolled more than 500,000 participants during 2006-2010 who were aged between 40-69 years and living in the United Kingdom.[27] Participants were identified from National Health Service (NHS) population registries. Eligible people lived within a reasonable distance (up to 10 miles radius) from an assessment center. A detailed lifestyle questionnaire, physical measurement, and blood, urine and saliva samples were collected at recruitment. Blood samples are stored at -80°C for the working archive and in liquid nitrogen vapour (-196°C) for the backup archive. Participants are followed up every 2-3 years with a questionnaire and collection of biological samples. Cancer diagnoses and deaths are identified by linkage to the Office for National Statistics (ONS) registry in England and Wales, and to the Registrar General's Office (RGO) registry in Scotland.

**VITAL: VITamins And Lifestyle Study**
The VITAL cohort enrolled 77,738 participants aged 50-76 during 2000-2002 in Washington state, USA.[28] The primary aim was to study the association between supplement use and cancer risk. The identification of participants was done through a commercial mailing list. At baseline, participants completed a detailed questionnaire including information on supplement use, diet, health history and risk factors. Three months after the baseline questionnaire, participants were asked to return a buccal cell sample through a DNA kit, and samples were stored at -80°C. Information on cancers was obtained by annual linkage to the Surveillance, Epidemiology, and End Results (SEER) file in western Washington state. In

addition, deaths were ascertained with an annual linkage to Washington State death files. Participants were censored when they moved out of the catchment area, based on information from the US Postal Service. Those with a history of lung cancer at baseline or who were subsequently diagnosed with rare lung cancer morphologies (e.g., lymphoma) were excluded from LC3 analyses.

**WHI: Women's Health Initiative**

WHI is a long-term health study that enrolled over 161,808 post-menopausal women from 40 centers across the US between 1993-1998.[29,30] The study (1993-2005) included three randomized controlled clinical trials (68,132 women) and an observational study (93,676 women), with all participants aged from 50 to 79 at enrollment. Women were identified from different sources such as motor vehicle registration lists, driver's license lists, health insurance companies, commercial mailing lists. Exclusion criteria included invasive cancer in the past 10 years, previous breast cancer, heart disease, obstructive lung disease with ventilation or oxygen, severe chronic liver disease, dialysis, sickle cell anemia, alcoholism, drug dependency, and mental illness. The Clinical Trials included a hormone therapy trial, a dietary modification trial, and a calcium/vitamin D trial. Two extension studies were made in 2005-2010 enrolling 115,400 women and in 2010-2020 involving 93,500 women with follow-up continuing through 2027. The baseline questionnaire included medical history, reproductive history, family history, personal habits, hormone use questions, and medications. In addition, physical measurements were recorded and a clinical exam was performed. Biospecimens were collected from fasting women at the baseline clinic visit and frozen at -70°C. Cancer diagnoses and deaths were centrally adjudicated, and SEER coded for the original WHI study and extensions. For our lung cancer analyses, we included women in the WHI observational study (OS) or the non-intervention arms of the clinical trial (CT). For a small number of participants, age at enrollment may differ from the age inclusion criteria due to participants requesting a correction to their birth date after enrollment. More information is available at www.whi.org.

**WHS: Women's Health Study**

The Women's Health Study (WHS) is a randomized, double-blind, placebo-controlled trial started in 1993 at Brigham and Women's Hospital and Harvard Medical School to study the benefits and risks of low-dose aspirin, vitamin-E, and beta-carotene in the prevention of cardiovascular disease and cancer in women.[31] Participants were recruited among members of health professional organizations and included women aged 45 or older, postmenopausal or with no intention of pregnancy, without history of heart disease or cancer (except non-melanoma skin cancer) or other serious disease, who were not taking aspirin, anti-inflammatory drugs, anticoagulants, corticosteroids, or supplements. At baseline, 39,876 women were enrolled and asked to complete a questionnaire including health history, dietary habits, and risk factors. A blood sample was obtained at baseline and stored at -170°C. Participants were randomized using a 2x2x2 factorial design into 4 equal groups: aspirin, vitamin E, beta-carotene or placebo. Cancer cases were self-reported and subsequently confirmed by medical records. Vital status was confirmed by medical record and a copy of the death certificate, and a linkage to the National Death Index was done at the end of the trial period for those who were lost to follow-up. We included all trial arms of WHS in the LC3.

### Descriptions of LDCT screening studies participating in the INTEGRAL Nodule Malignancy project

**PanCan: Pan-Canadian Early Detection of Lung Cancer Study**
This study prospectively recruited 2537 subjects between Sept 24, 2008, and Dec 17, 2010.[32] Participants had at least a 2% 6-year risk of lung cancer as estimated by the PanCan model, a precursor to the validated PLCOm2012 model. The eligible subjects were ever smokers, aged 50-75 years old who had no previous history of invasive cancer. The subjects were recruited at eight centers across Canada (Vancouver, Calgary, Hamilton, Toronto, Ottawa, Quebec, Halifax, and St. John's. All study participants completed a detailed questionnaire, spirometry, collection of blood specimens for biomarker measurement and LDCT at baseline. Participants with an abnormal LDCT results (the presence of any noncalcified non-perifissural pulmonary nodule or a nodule with non-solid area of at least 1mm diameter) were followed up every 3-24 months. A nodule was considered benign if it showed a benign calcification pattern (eg, fully calcified, popcorn calcification) or if the size and density of was unchanged for longer than 2 years.

**UKLS: The UK Lung Cancer Pilot Screening Trial**
The UKLS is a randomized controlled trial of LDCT screening in individuals aged 50-75 years with at least 4.5% risk of developing lung cancer in 5 years based on the Liverpool Lung Project (LLPv2) risk model.[33,34] The subjects with any comorbidity that unequivocally contraindicated either screening or treatment were excluded. There were 4055 individuals recruited in two geographical areas, covering six primary care trusts around Liverpool and Cambridge, from 2011 to 2013. The participants were randomized to receive either LDCT or usual care, 1994 of which underwent LDCT scans. Most participants had their lung function measured, provided blood samples and completed detailed epidemiological, psychosocial and health economics questionnaires at baseline. The LDCT subjects received a baseline scan and nodules were assessed by two local radiologists and placed in one of four pre-defined nodule categories: Category 4 (Solid: volume >500mm$^3$ or diameter >10mm; Part solid: solid component- volume >500mm$^3$); Category 3 (Solid: volume 50 - 500mm$^3$ or diameter 5 - 9.9mm; Part solid: non-solid component diameter >5mm, solid component :15 - 500mm$^3$ or diameter 3 - 9.9mm; Non-solid: diameter ≥ 5mm), Category 2 (Solid: volume 15 - 49mm$^3$ or diameter 3 -4.9mm; Part solid: solid component volume <15mm$^3$ or diameter < 3mm; Non solid diameter 3 - 4.9mm); Category 1 (Benign nodule or volume <15mm$^3$ or diameter <3mm or features of an intrapulmonary lymph node). Consensus nodule category was assigned following central reading at the Royal Brompton Hospital, with a read for arbitration when necessary. Category 4 nodules were immediately referred for work-up and clinical management. Category 3 nodules identified in the baseline scan were re-analyzed in follow-up CT scans at three and twelve months; Category 2 nodules at twelve months only. Category 1 nodules were considered normal and had no further follow-up. Growth of nodules was based on their characteristics and volume doubling time (VDT); i.e. VDT <400 days or new solid component of non-solid nodule was classified as growth in the UKLS trial and were referred to the trial participating MDT clinic for work-up and clinical management.

**IELCAP-Toronto: the International Early Lung Cancer Action Program – Toronto**
This single arm early detection of lung cancer research study at Princess Margaret Cancer Centre, Toronto, Canada joined the international Early Lung Cancer Action Program (I-ELCAP) in 2003.[35–37] Ever smokers of more than 10 pack-years, aged 50 years or older were eligible for LDCT screening. A total of 4782 individuals have been enrolled in the Greater Toronto Area, and 4161 of those have the clinic-epidemiological data needed to be part of INTEGRAL. Participants were administered a LDCT scan along with a standard study

questionnaire at baseline. Blood samples were systematically collected at baseline since 2006. In this study, a LDCT results was considered to be normal if no nodules of the size of 5 mm or larger was found. In case of a positive LDCT result, an indeterminate noncalcified solid or part-solid nodule 5 mm or larger, or a nonsolid nodule 8 mm or larger was detected. The abnormal scans were followed up at 1-2 years depending on nodule size and shape. The location, size, consistency, calcifications and spiculations were determined for each nodule.

**P-IELCAP: the Pamplona International Early Lung Cancer Action Program**
P-IELCAP is an ongoing lung cancer screening program in Spain which began recruitment in September 2000.[38] It is part of the I-ELCAP consortium. As of December 2019, the program had enrolled 3825 asymptomatic subjects older than 40 years of age, including former of current smokers (≥10 pack-years), cancer-free 5 years prior to enrollment. The subjects completed LDCT, spirometry, blood collection and a health questionnaire at baseline and during follow up. Subjects with a positive baseline LDCT (noncalcified nodules ≥5 or 6 mm depending on the year of enrollment), had imaging follow up at 1-3 months. Subjects with negative scans or small pulmonary nodules at baseline had follow up LDCTs at annual intervals.

**NLST: The National Lung Screening Trial**
The NLST is a randomized trial which is a merger of two studies: the NCI Lung Screening Study (LSS) and the American College of Radiology Imaging Network (ACRIN).[39] The NLST recruited 53,454 subjects from 2002 to 2004 and followed them up by 2009. The participants were randomly assigned to two screening groups of LDCT and chest radiography with almost identical sizes. The eligible participants were 55 to 74 years old subjects who were either heavy current smokers (at least 30 pack-years) or former smokers who quit smoking within the last 15 years. The subjects underwent three screenings at 1-year intervals if they were not diagnosed for lung cancer in the previous visits. At each screening visit, the participants were required to complete a vital status questionnaire, have their blood, sputum, and urine collected, and underwent LDCT or chest radiography. All screening examinations were performed in accordance with a standard protocol. The LDCT scans were classified as abnormal if any noncalcified nodule measuring at least 4 mm in any diameter was present. Any noncalcified nodule or mass in radiographic images also indicated an abnormality.

**PLuSS: The Pittsburgh Lung Screening Study**
PLuSS is a community-based research cohort of current and former smokers, screened with LDCT and followed for lung cancer.[40–42] Between 2002 and 2005, PLuSS enrolled 3,642 study participants. Subjects were eligible if they were 50 to 79 years old, had no personal history of lung cancer, and were current or former smokers with at least half-pack per day for at least 25 years, and if quit smoking, no more than 10 years prior to enrollment. Upon study entry, participants completed a questionnaire and underwent CT screening and spirometry. Approximately one year later, participants underwent repeat CT screening. All PLuSS participants are followed for lung cancer-related endpoints but 977 PLuSS subjects judged to be at the highest risk for lung cancer were also recruited to an extension of the study. Subjects in PLuSS-extension underwent LDCT screening every other year and spirometry every year (through 2015). Non-calcified nodules identified by CT were categorized into those with low, moderate or high lung cancer suspicion rating, based on mean diameter, spiculation, timing of first appearance, and subjective criteria from the interpreting radiologist. Participants with moderate or high lung cancer suspicion were referred to physician-directed diagnostic follow-up. Those with low lung cancer suspicion were recommended annual repeat CT screening if they had micronodules (<4 mm) only, and 6-month CT follow-up if they had larger (≥4 mm) nodules. Biospecimen were collected at

enrollment and, from those in PLuSS-extension, during yearly study visits for biomarker and risk prediction studies.

#### References (Supplement)


1. Schatzkin A, Subar AF, Thompson FE, Harlan LC, Tangrea J, Hollenbeck AR, et al. Design and serendipity in establishing a large cohort with wide dietary intake distributions: the National Institutes of Health-American Association of Retired Persons Diet and Health Study. Am J Epidemiol. 2001 Dec 15;154(12):1119–25.
2. The ATBC Cancer Prevention Study Group. The alpha-tocopherol, beta-carotene lung cancer prevention study: Design, methods, participant characteristics, and compliance. Ann Epidemiol. 1994 Jan;4(1):1–10.
3. Johns Hopkins Bloomberg School of Public Health. George W. Comstock Center for Public Health Research and Prevention: CLUE studies. [Internet]. [cited 2022 Feb 21]. Available from: https://www.jhsph.edu/research/centers-and-institutes/george-w-comstock-center-for-public-health-research-and-prevention/clue_research_activities.html
4. Calle EE, Rodriguez C, Jacobs EJ, Almon ML, Chao A, McCullough ML, et al. The American Cancer Society Cancer Prevention Study II Nutrition Cohort. Cancer. 2002 May 1;94(9):2490–501.
5. Rohan TE, Soskolne CL, Carroll KK, Kreiger N. The Canadian Study of Diet, Lifestyle, and Health: design and characteristics of a new cohort study of cancer risk. Cancer Detect Prev. 2007;31(1):12–7.
6. Riboli E, Kaaks R. The EPIC Project: rationale and study design. European Prospective Investigation into Cancer and Nutrition. Int J Epidemiol. 1997;26 Suppl 1:S6-14.
7. Riboli E, Hunt KJ, Slimani N, Ferrari P, Norat T, Fahey M, et al. European Prospective Investigation into Cancer and Nutrition (EPIC): study populations and data collection. Public Health Nutr. 2002 Dec;5(6B):1113–24.
8. Slimani N, Kaaks R, Ferrari P, Casagrande C, Clavel-Chapelon F, Lotze G, et al. European Prospective Investigation into Cancer and Nutrition (EPIC) calibration study: rationale, design and population characteristics. Public Health Nutr. 2002 Dec;5(6B):1125–45.
9. Pourshams A, Khademi H, Malekshah AF, Islami F, Nouraei M, Sadjadi AR, et al. Cohort Profile: The Golestan Cohort Study--a prospective study of oesophageal cancer in northern Iran. Int J Epidemiol. 2010 Feb;39(1):52–9.
10. Swerdlow AJ, Jones ME, Schoemaker MJ, Hemming J, Thomas D, Williamson J, et al. The Breakthrough Generations Study: design of a long-term UK cohort study to investigate breast cancer aetiology. Br J Cancer. 2011/09/06. 2011 Sep 27;105(7):911–7.
11. Giovannucci E, Ascherio A, Rimm EB, Colditz GA, Stampfer MJ, Willett WC. Physical activity, obesity, and risk for colon cancer and adenoma in men. Ann Intern Med. 1995 Mar 1;122(5):327–34.
12. Krokstad S, Langhammer A, Hveem K, Holmen T, Midthjell K, Stene T, et al. Cohort Profile: The HUNT Study, Norway. Int J Epidemiol. 2013 Aug 1;42(4):968–77.
13. Milne RL, Fletcher AS, MacInnis RJ, Hodge AM, Hopkins AH, Bassett JK, et al. Cohort Profile: The Melbourne Collaborative Cohort Study (Health 2020). Int J Epidemiol. 2017 Dec 1;46(6):1757-1757i.
14. Belanger CF, Hennekens CH, Rosner B, Speizer FE. The Nurses' Health Study. Am J Nurs. 1978 Jun;78(6):1039–40.
15. Hallmans G, Ågren Å, Johansson G, Johansson A, Stegmayr B, Jansson J-H, et al. Cardiovascular disease and diabetes in the Northern Sweden Health and Disease Study Cohort- evaluation of risk factors and their interactions. Scand J Public Health. 2003 Nov 25;31(61_suppl):18–24.
16. Norberg M, Wall S, Boman K, Weinehall L. The Västerbotten Intervention

Programme: background, design and implications. Glob Health Action. 2010 Dec 22;3(1):4643.
17. Toniolo PG, Pasternack BS, Shore RE, Sonnenschein E, Koenig KL, Rosenberg C, et al. Endogenous hormones and breast cancer: a prospective cohort study. Breast Cancer Res Treat. 1991 May;18 Suppl 1:S23-6.
18. Toniolo PG, Levitz M, Zeleniuch-Jacquotte A, Banerjee S, Koenig KL, Shore RE, et al. A prospective study of endogenous estrogens and breast cancer in postmenopausal women. J Natl Cancer Inst. 1995 Feb 1;87(3):190–7.
19. Steering Committee of the Physicians' Health Study Research Group. Final report on the aspirin component of the ongoing Physicians' Health Study. N Engl J Med. 1989;321(3):129–35.
20. Prorok PC, Andriole GL, Bresalier RS, Buys SS, Chia D, Crawford ED, et al. Design of the Prostate, Lung, Colorectal and Ovarian (PLCO) Cancer Screening Trial. Control Clin Trials. 2000 Dec;21(6 Suppl):273S-309S.
21. Oken MM, Hocking WG, Kvale PA, Andriole GL, Buys SS, Church TR, et al. Screening by chest radiograph and lung cancer mortality: the Prostate, Lung, Colorectal, and Ovarian (PLCO) randomized trial. J Am Med Assoc. 2011 Nov 2;306(17):1865–73.
22. Signorello LB, Hargreaves MK, Steinwandel MD, Zheng W, Cai Q, Schlundt DG, et al. Southern Community Cohort Study: establishing a cohort to investigate health disparities. J Natl Med Assoc. 2005 Jul;97(7):972–9.
23. Hankin JH, Stram DO, Arakawa K, Park S, Low SH, Lee HP, et al. Singapore Chinese Health Study: development, validation, and calibration of the quantitative food frequency questionnaire. Nutr Cancer. 2001;39(2):187–95.
24. Yuan JM, Ross RK, Wang XL, Gao YT, Henderson BE, Yu MC. Morbidity and mortality in relation to cigarette smoking in Shanghai, China. A prospective male cohort study. JAMA. 1996 Jun;275(21):1646–50.
25. Shu X-O, Li H, Yang G, Gao J, Cai H, Takata Y, et al. Cohort Profile: The Shanghai Men's Health Study. Int J Epidemiol. 2015 Jun;44(3):810–8.
26. Zheng W, Chow W-H, Yang G, Jin F, Rothman N, Blair A, et al. The Shanghai Women's Health Study: rationale, study design, and baseline characteristics. Am J Epidemiol. 2005 Dec;162(11):1123–31.
27. Sudlow C, Gallacher J, Allen N, Beral V, Burton P, Danesh J, et al. UK Biobank: an open access resource for identifying the causes of a wide range of complex diseases of middle and old age. PLoS Med. 2015 Mar 31;12(3):e1001779–e1001779.
28. White E, Patterson RE, Kristal AR, Thornquist M, King I, Shattuck AL, et al. VITamins And Lifestyle cohort study: study design and characteristics of supplement users. Am J Epidemiol. 2004 Jan 1;159(1):83–93.
29. Langer RD, White E, Lewis CE, Kotchen JM, Hendrix SL, Trevisan M. The Women's Health Initiative Observational Study: baseline characteristics of participants and reliability of baseline measures. Ann Epidemiol. 2003 Oct;13(9 Suppl):S107-21.
30. The Women's Health Initiative Study Group. Design of the Women's Health Initiative clinical trial and observational study. The Women's Health Initiative Study Group. Control Clin Trials. 1998 Feb;19(1):61–109.
31. Buring JE and Hennekens CH for the Women's Health Study Research Group. The Women's Health Study: Rationale and background. J Myocard Ischemia. 1992;4:30–40.
32. Tammemagi MC, Schmidt H, Martel S, McWilliams A, Goffin JR, Johnston MR, et al. Participant selection for lung cancer screening by risk modelling (the Pan-Canadian Early Detection of Lung Cancer [PanCan] study): a single-arm, prospective study. Lancet Oncol. 2017 Oct 16;18(11):1523–31.
33. Field JK, Duffy SW, Baldwin DR, Whynes DK, Devaraj A, Brain KE, et al. UK Lung

Cancer RCT Pilot Screening Trial: baseline findings from the screening arm provide evidence for the potential implementation of lung cancer screening. Thorax. 2016 Feb;71(2):161–70.
34. Field JK, Duffy SW, Baldwin DR, Brain KE, Devaraj A, Eisen T, et al. The UK Lung Cancer Screening Trial: a pilot randomised controlled trial of low-dose computed tomography screening for the early detection of lung cancer. Health Technol Assess. 2016;20(40):1–146.
35. Roberts HC, Patsios D, Paul NS, McGregor M, Weisbrod G, Chung T, et al. Lung cancer screening with low-dose computed tomography: Canadian experience. Can Assoc Radiol J. 2007 Oct;58(4):225–35.
36. Kavanagh J, Liu G, Menezes R, O'Kane GM, McGregor M, Tsao M, et al. Importance of long-term low-dose CT follow-up after negative findings at previous lung cancer screening. Radiology. 2018 Jul;180053.
37. Aggarwal R, Lam ACL, McGregor M, Menezes R, Hueniken K, Tateishi H, et al. Outcomes of long-term interval rescreening with low-dose computed tomography for lung cancer in different risk cohorts. J Thorac Oncol. 2019 Jun;14(6):1003–11.
38. Mesa-Guzmán M, González J, Alcaide AB, Bertó J, De-Torres JP, Campo A, et al. Surgical outcomes in a lung cancer screening program using low dose computed tomography. Arch Bronconeumol. 2021 Feb;57(2):101–6.
39. National Lung Screening Trial Research Team, Aberle DR, Adams AM, Berg CD, Black WC, Clapp JD, et al. Reduced lung-cancer mortality with low-dose computed tomographic screening. N Engl J Med. 2011 Aug 4;365(5):395–409.
40. Wilson DO, Weissfeld JL, Fuhrman CR, Fisher SN, Balogh P, Landreneau RJ, et al. The Pittsburgh Lung Screening Study (PLuSS): outcomes within 3 years of a first computed tomography scan. Am J Respir Crit Care Med. 2008 Nov;178(9):956–61.
41. Weissfeld JL, Lin Y, Lin H-M, Kurland BF, Wilson DO, Fuhrman CR, et al. Lung cancer risk prediction using common SNPs located in GWAS-identified susceptibility regions. J Thorac Oncol. 2015 Nov;10(11):1538–45.
42. Leng S, Diergaarde B, Picchi MA, Wilson DO, Gilliland FD, Yuan J-M, et al. Gene promoter hypermethylation detected in sputum predicts FEV(1) decline and all-cause mortality in smokers. Am J Respir Crit Care Med. 2018 Jul;198(2):187–96.
